# Performance assessment of an electrostatic filter-diverter stent cerebrovascular protection device

**DOI:** 10.1101/2023.03.31.23288032

**Authors:** Beatriz Eguzkitza, David Oks, José A. Navia, Guillaume Houzeaux, Constantine Butakoff, María Fisa, Ariadna Campoy Millán, Mariano Vázquez

## Abstract

Stroke is the second leading cause of death worldwide. Nearly two-thirds of strokes are produced by cardioembolisms, and half of cardioembolic strokes are triggered by Atrial Fibrillation (AF), the most common type of arrhythmia. A more recent cause of cardioembolisms is Transcatheter Aortic Valve Replacements (TAVRs), which may onset post-procedural adverse events such as stroke and Silent Brain Infarcts (SBIs), for which no definitive treatment exists, and which will only get worse as TAVRs are implanted in younger and lower risk patients. It is well known that some specific characteristics of elderly patients may lower the safety and efficacy of anticoagulation therapy, making it a real urgency to find alternative therapies. The device introduced in this paper offers an anticoagulant-free method to prevent stroke and SBIs, imperative given the growing population of AF and elderly patients. This work analyzes a design based on a patented medical device, intended to block cardioembolisms from entering the cerebrovascular system, with a particular focus on AF, and potentially TAVR patients. The study has been carried out in two stages. Both of them use computational fluid dynamics (CFD) coupled with Lagrangian particle tracking to analyse the efficacy of a novel patented medical device intended to block cardioembolisms from entering the cerebrovascular system, with a particular focus on AF, and potentially TAVR patients. The studied device consists of a strut structure deployed at the base of the treated artery. Particles of different sizes are used to model dislodged debris, which could potentially lead to cerebral embolisms if transported into these arteries.

The first stage of the work evaluates a variety of strut thicknesses and inter-strut spacings, contrasting with the device-free baseline geometry. The analysis is carried out by imposing flowrate waveforms characteristic of both healthy and AF patients. Boundary conditions are calibrated to reproduce physiological flowrates and pressures in a patient’s aortic arch. Results from numerical simulations indicate that the device blocks particles of sizes larger than the inter-strut spacing. It was found that lateral strut space had the highest impact on efficacy.

In the second stage, the optimal geometric design from the first stage was employed, with the addition of lateral struts to prevent the filtration of particles and electronegatively charged strut surfaces, studying the effect of electrical forces on the clots if they are considered charged. Flowrate boundary conditions were used to emulate both healthy and AF conditions. When deploying the electronegatively charged device in all three aortic arch arteries, the number of particles entering these arteries was reduced on average by 62.6% and 51.2%, for the healthy and diseased models respectively, matching or surpassing current oral anticoagulant efficacy. The device demonstrated a two-fold mechanism for filtering emboli: while the smallest particles are deflected by electrostatic repulsion, avoiding microembolisms, which could lead to cognitive impairment, the largest ones are mechanically filtered since they cannot fit in between the struts, effectively blocking the full range of particle sizes analyzed in this study.

## 1 Introduction

Stroke is the second leading cause of death worldwide [1]. Nearly two-thirds of strokes are produced by cardioembolisms [2]. Cardioembolic strokes are caused by blood clots that form in the heart, due to disease (e.g., atrial fibrillation) or a cardiac intervention (e.g., transcatheter aortic valve replacements or left atrial appendage occluders), and travel through the bloodstream into the brain. Half of the cardioembolic strokes are caused by Atrial Fibrillation (AF), a heart condition that causes an irregular and often abnormally fast heart rate with a significant reduction in the cardiac output. It is proven that this dysfunction increases the propensity of emboli, which tend to travel through the Brachiocephalic Trunk (BCT), Left Carotid Common Artery (LCCA), and Left Subclavian Artery (LSA), to the brain, obstructing the superior arteries and triggering cerebral strokes [3].

### 1.1 Why is atrial fibrillation a critical problem?

AF is the most common heart rhythm disorder, responsible for approximately one-third of hospitalizations for cardiac rhythm disturbances in the US. The prevalence and incidence of AF are increasing. It is predicted to affect 6-12 million people in the US by 2050 and 17.9 million in Europe by 2060, significantly impacting wellbeing and healthcare costs. AF is associated with increased morbidity and mortality, due to the risk of ischemic stroke, systemic embolism, heart failure, and cognitive impairment, reducing the quality and quantity of life in these patients [4]. This condition is associated with a six-fold increase in stroke. Moreover, patients with previous ischemic stroke are at an even higher risk [5]. In the case of cardioembolic strokes (two-thirds of the total), echocardiographic and pathologic studies suggest that when a source can be identified, approximately 90% of such strokes can be attributed to thrombus formation in the left atrial appendage [2].

### 1.2 What are the existing treatments for stroke and what are their drawbacks?

The primary approach to prevent ischaemic stroke risk associated with AF, are anticoagulation therapies, which have a proven efficacy. At least four large clinical trials have clearly demonstrated that anticoagulation with warfarin decreases the risk of stroke by 50-80% [6, 7, 8, 9]. In relatively recent trials, the newer oral anticoagulants (OACs), such as dabigatran, rivaroxaban, apixaban, and edoxaban, have proven to be similarly effective to warfarin for the prevention of stroke and thromboembolism [10]. However, anticoagulants have significant drawbacks. Although anticoagulants reduce 30-day mortality from ischaemic stroke, these agents increase intracranial haemorrhage-related mortality [11]. Moreover, patients having a stroke despite being on therapy with an oral anticoagulant are at high risk of recurrent ischaemic strokes [12]. In the ANAFIE registry, patients at high bleeding risk had higher incidences of stroke, major bleeding, intracranial haemorrhage, gastrointestinal bleeding, cardiovascular events, and all-cause death than patients in the reference group, despite a high prevalence of OAC therapy (89.0%) [13]. In elderly patients, non-adherence to OAC treatment, associated commorbidities and additional risk factors can significantly increase the incidence and severity of cerebrovascular accidents. In this age group, a delicate balance may exist between multiple conditions, being thrombotic disease, chronic kidney disease, cancer, coronary artery disease, and heart failure, some of the most challenging scenarios encountered in clinical practice [13]. In addition, OACs have multiple contraindications [14]:

- prior intracranial haemorrhage or diseases predisposing to intracranial haemorrhage,
- active gastrointestinal bleeding or diseases predisposing to gastrointestinal bleeding (such as active ulcer), or inflammation of the gastrointestinal tract,
- anaemia, defined as Hgb level below 8 mg dl,
- thrombocytopenia, defined as PLT *<* 50, 000 platelets per μL,
- end-stage liver disease, and
- allergy

Decisions to start effective AF-related stroke thromboprophylaxis following an acute ischaemic stroke or intracerebral haemorrhage are rarely clear-cut: patients have reluctance and own prejudices, and relative contraindications are influenced by their individual clinicians’ perceptions of risks and benefits [5]. It is therefore necessary to rigorously evaluate the current status of oral anticoagulation agent use for AF-related stroke prevention. Considering the difficulties involved in anticoagulant treatments, it is only reasonable to devise CEPDs capable of matching or surpassing current OAC efficacy.

### 1.3 What is the relation between TAVR and stroke?

TAVR has emerged as an alternative, rapidly evolving non-invasive procedure for patients with severe aortic stenosis and medium-to-high surgical risk. By 2025, there will be an estimated 280,000 TAVR procedures performed worldwide and the total market will exceed $8 billion. Although this highly promising treatment modality results in less morbidity, shorter time to recovery and similar mortality rates, it is still associated with one of the most devastating and feared complications: cerebral embolism, which in turn may cause stroke [15]. Stroke is associated with a 6-fold increase in mortality in TAVR cohorts, a moderate to severe permanent disability in up to 40% of survivors [5], a 4.7-fold increased risk of permanent work disability [2], social isolation and significant financial strain in 80% of stroke survivors [11], and an increased risk of readmission in patients with stroke after cardiac catheterization [16].

The time in between the TAVR procedure and the cardioembolic event is an important factor when choosing stroke prevention treatments. Most of them occur in the acute phase following TAVR where cerebral embolic events are frequent [17]. Nonetheless, according to the ADVANCE trial, half of the reported strokes occurred between day 2 and 30 after the TAVR procedure [18]. Moreover, evidence is mounting on ischaemic brain lesions being produced after day 30, causing SBIs and long-term neurological symptoms. New ischaemic brain lesions were found in 74% to 100% of patients on diffusion-weighted magnetic resonance imaging (DW-MRI) after TAVR [19]. Although studies have shown that SBI may not be related to apparent short-term neurological symptoms, evidence points to an association with accelerated cognitive decline and strengthening of the risk of long-term dementia (most commonly, Alzheimer’s disease) [20]. Later events are associated with patient specific factors [21]. SBIs have been associated with accelerated cognitive decline and higher risk of long-term dementia. The situation is especially worrying given that TAVRs are being implanted in increasingly younger and lower risk patients, hence potentially increasing the prevalence of dementia.

### 1.4 What is the state of the art of cerebral protection devices?

As mentioned, in patients undergoing TAVR, stroke remains a potentially devastating complication associated with significant morbidity and mortality. This is especially worrying, given that TAVR is now indicated in aortic stenosis low- and intermediate-risk patients [22], and that stroke rate 30 days post-TAVR has been reported at 3.4% in low risk-patients [23]. To prevent debris from embolizing to the brain during the procedure and reduce the risk of stroke, cerebroembolic protection devices (CEPDs) were developed [24]. These devices are implanted during the procedure and up to ≥ 2 days after, since the clogging of CEPDs impedes them from being used to prevent ischaemic strokes *>* 2 days post-TAVR. Nonetheless, as explained above, risk of stroke may not be limited to the procedure itself or the perioperative period. Moreover, the clinical benefit of current CEPDs in reducing strokes, transient ischaemic attacks or death remains unknown [25, 26, 27]. It is therefore worrying that for strokes triggered by AF or other conditions extended in time, there are no currently approved CEPD in the market. The SENTINEL-LIR study demonstrated that embolic debris captured by the SENTINEL-CPS (Cerebral Protection System) during TAVR in low- to intermediate-risk patients was similar to that in previous studies conducted among higher-risk patients. These findings suggest lower-risk patients undergoing TAVR have potentially a similar embolic risk as high-risk patients, as evidenced by embolic debris capture [28]. Most captured debris had a size of *<* 500 μm, with 78% between 150 and 500 μm. Larger size particles (≥1000 μm), which can cause significant vessel obstruction, were present in 67% of cases. Therefore, the Sentinel CEPD can functionally capture large debris that may cause a severe stroke. In contrast, debris on the micrometer scale may pass through the gaps between the filters and arteries, leading to stroke even in CEPD-protected territories (with less likelihood of severe symptoms) [29]. Dedicated meta-analyses demonstrated that SBIs of small volume 3 mm^3^ are independent predictors of later stroke and mortality [16], further highlighting the need for a CEPD able to filter small debris in the long term post-TAVR. Such a device would be a radical improvement for a large population at risk.

### 1.5 How can this paper help improve the current situation?

The main purpose of the work presented in this paper can be described with the simple motto: ***Save the brain***. The proposed solution consists of a stent attached to an electronegatively charged strut structure, intended to block the passage of clots or deflect their trajectories thanks to both fluid- and electro-dynamical forces on the clots. This can be achieved thanks to the electrostatic repulsion acting on naturally electronegatively charged blood clots, and its geometrical design, capable of filtering thrombi based on the distance between struts. The main objective of this device is to reduce or eliminate the percentage of blood clots entering the BCT, LCCA, and LSA, while maintaining at least 80% of the natural delivery of oxygenated blood to the brain. The Section 4.3 will demonstrate that the presence of the devices in all three arteries reduces the flowrate in a maximum of 7.5% in the case of BCT, and in less than 2% in the other two aortic arch arteries. This paper provides the details of the work carried out using Computational Fluid Dynamics (CFD) coupled with Lagrangian particle transport to validate the efficacy of the deflecting medical device positioned at the base of the BCT, LCCA, and LSA. A sufficiently large number of particles were injected into the domain in order to assure significant statistical samples for validating the particle-deflecting capabilities of the device. The study is divided in two main stages. The first one analyzes the effect of the thickness and shape of the strut design on the device performance. In the first stage, the purely hydrodynamic effect of the device is analyzed using a CFD and a particle transport model. The device is placed at the root of the LCCA and the optimal strut thickness is identified by analyzing the trajectories of particles suspended in the flow. The analysis showed a low efficacy for the deflection of thrombi and identified a deficiency in the initial design which is was letting particles pass through the lateral struts. To overcome this deficiency, extra struts are added in the second design employed in the second stage of the work, oriented perpendicularly to the original struts. The second stage of this project, proposes to simulate struts that are electrically charged on their surface. Considering that blood clots are negatively charged, the strut surface would be negatively charged too in order to repel the clots. To achieve the negative charge of the struts, a proposed solution is to coat the Nitinol struts with a graphene oxide coating, with appropriate electrostatic properties. Due to the latter, it was possible to deflect even the smallest particles from entering the aortic arch arteries, offering an anticoagulant-free method to prevent SBIs.

## 2 Methods

### 2.1 Aortic arch geometry

The aortic arch geometry used for computational simulations was designed based on an anatomical model from a previous study [30]. In order to adapt the geometry to the requirements of the present study, some modifications were made using ANSA v22.1.0 (Beta Simulation Solutions, Lucerne, Switzerland). The applied modifications preserve the original structure of the aortic arch geometry, only in the LCCA and BCT significant changes (*i*.*e*., go from an anatomical structure to a synthetic one) have been applied. It was observed that the original geometry was not representative of an average BCT geometry, so it was manually corrected to adapt it to a more realistic one. Likewise, it was noticed that the LCCA presented a broader morphology at the entrance of the vessel, which could imply obtaining wrong conclusions, thus, it had to be corrected. Finally, let us comment that the aortic root was truncated to focus on the vessels in the aortic arch. For the inlet, the area corresponding to the left ventricle outflow tract (LVOT), with the coronary arteries and the aortic valve was replaced by a rigid tube to ensure the correct stabilisation of the fluid. At the outlet, the area corresponding to superior mesenteric, illiac, and renal arteries was dismissed. The resulting geometry used in this study can be observed in Figure 1.

**Figure 1:**
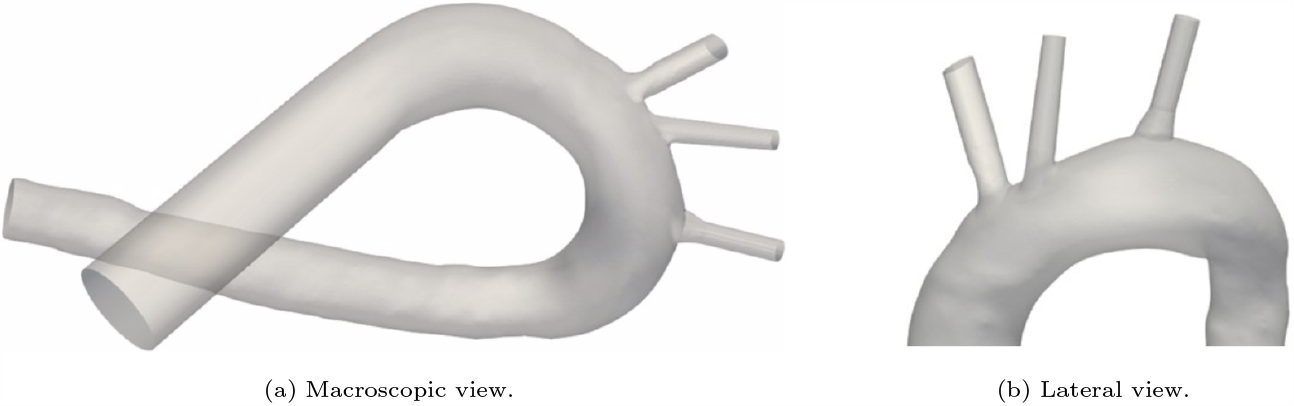
Aortic arch geometry. In the original geometry, the main vessels were segmented manually and recreated based on a CT scan of a cadaver. (a) Left ventricle, aortic valve and coronary arteries have been replaced in the inlet by a rigid tube, and superior mesenteric, iliac and renal arteries have been dismissed at the outlet. (b) Extrusion extensions have been applied at the outlet of the vessels to stabilise the simulated flux.

For the second stage of the present work, the device is implanted at the base of the three arteries: BCT, LCCA, and LSA. Figure 2 shows the patient-specific geometry of the aortic arch employed, with the arteries where the device will be placed. The inlet and outlet boundaries are labelled as:

- Inlet: Aortic root
- Outlets:
  - Brachiocephalic Trunk (BCT),
  - Left Carotid Common Artery (LCCA),
  - Left Subclavian Artery (LSA), and
  - Descending Aorta (DAO).

**Figure 2:**
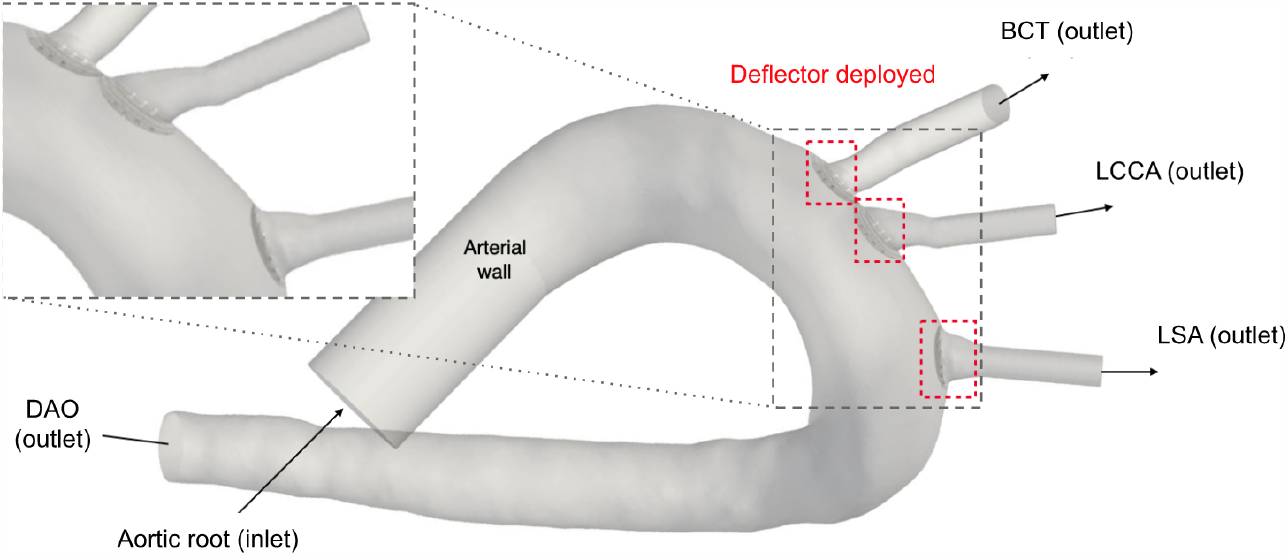
Geometry of the aortic arch and zoom-in of the deflectors deployed at all three arteries.

### 2.2 Device design

#### First stage

The emboli released from the aortic root due to TAVR-related debris tend to travel through the BCT, LCCA and LSA, obstructing the superior arteries and producing cerebral strokes, with the aforementioned associated consequences. Therefore, it is crucial that the introduced CEPD can be implanted in all three arteries of the aortic arch. The proposed solution, detailed in the patent no. US9517148 [31], fulfils exactly this requirement. It consists of a stent attached to a strut structure to block or deflect clot trajectories, as can be seen in Figure 4. This can be achieved thanks to its design, capable of filtering thrombi based on the distance between struts and eventually to modify trajectories of the smallest particles, responsible for SBI.

**Figure 3:**
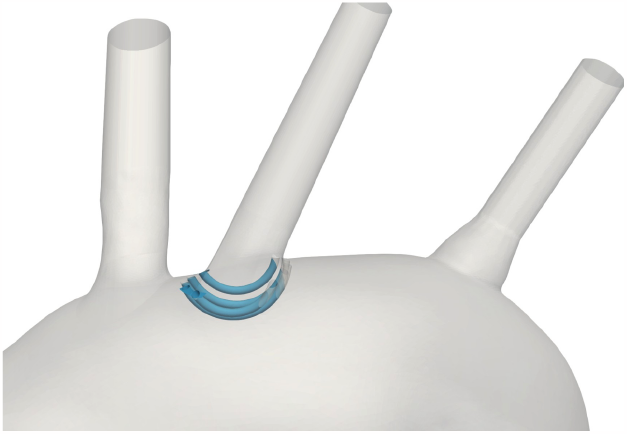
Deflector device (blue) implanted at the base of the LCCA.

**Figure 4:**
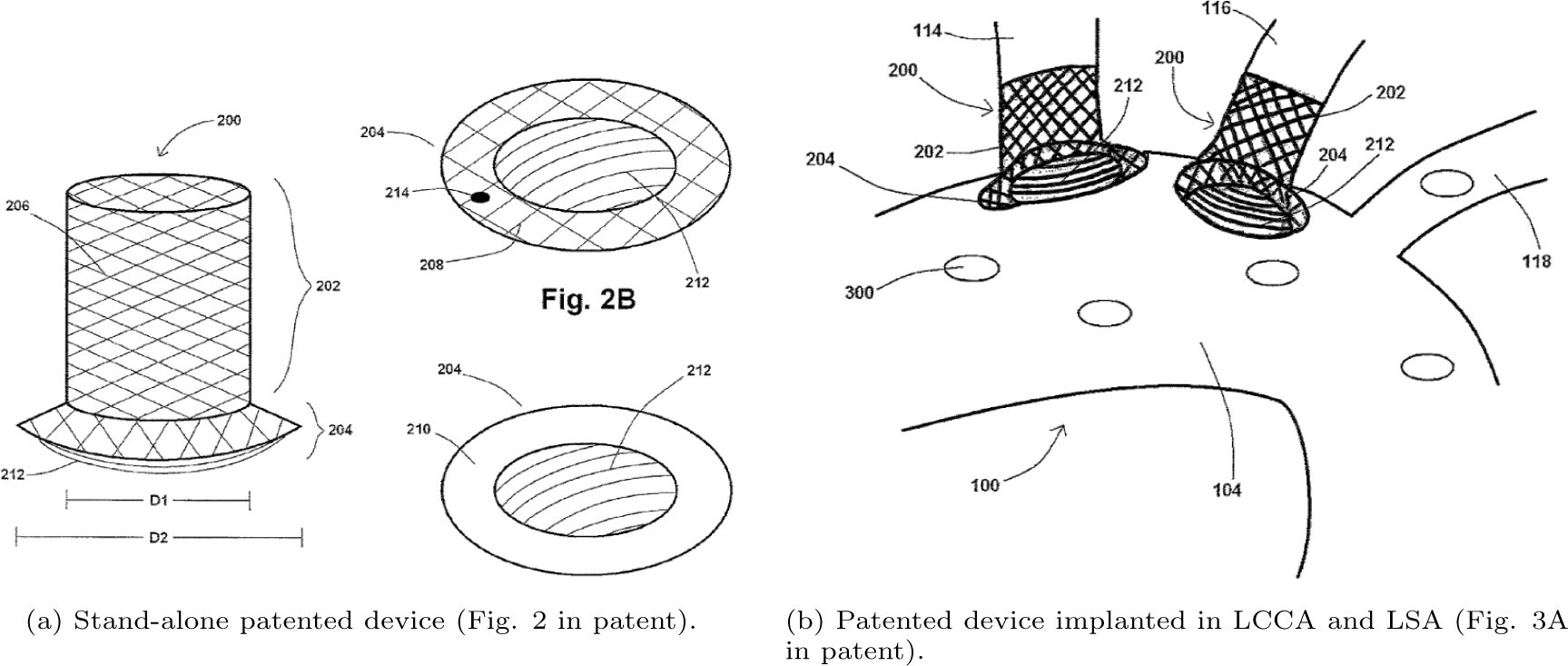
Diagrams from the patent of thrombus deflecting device, patent no. US9517148 [31].

It is worth noting that the patent provides some flexibility in design parameters such as the strut orientation, convexity, profile, and quantity. The main application of this device is to reduce or eliminate the percentage of embolisms entering the BCT, LCCA and LSA in the mid- to longterm in AF and TAVR patients, while maintaining the delivery of oxygenated blood to the brain. The introduced device has the objective of reducing the number of particles travelling to the brain through the branches of the aortic arch. To do so, in the first part of the study, it is implanted at the base of LCCA. Figure 3 shows the deflector device positioned at the LCCA.

The curved struts of the device are aligned with the aortic blood flow to effectively deflect particles from entering these vessels, without significantly reducing the fraction of ejected flow into the vessel where it is placed. The deflector structure has been designed with FreeCAD 0.19 [32]. For this step, both the dimensions stated in the reference study [33] and in the device patent have been considered. Five different deflector geometries have been generated considering two geometric parameters: the distance between deflector struts or strut interval (*L*_*si*_) and the strut thickness (*L*_*st*_). Figure 5a shows the deflector design and the above mentioned parameters. The resulting geometry dimensions are illustrated in Table 1. Figure 5b shows the aforementioned deflector geometries created with FreeCAD. As shown in Figure 3, in this first stage of the study the deflector device has been implanted at the LCCA. The choice to begin with this configuration takes into account different studies [34, 35] which suggest that cerebrovascular diseases, such as strokes, are significantly influenced by clots travelling through this artery.

**Figure 5:**
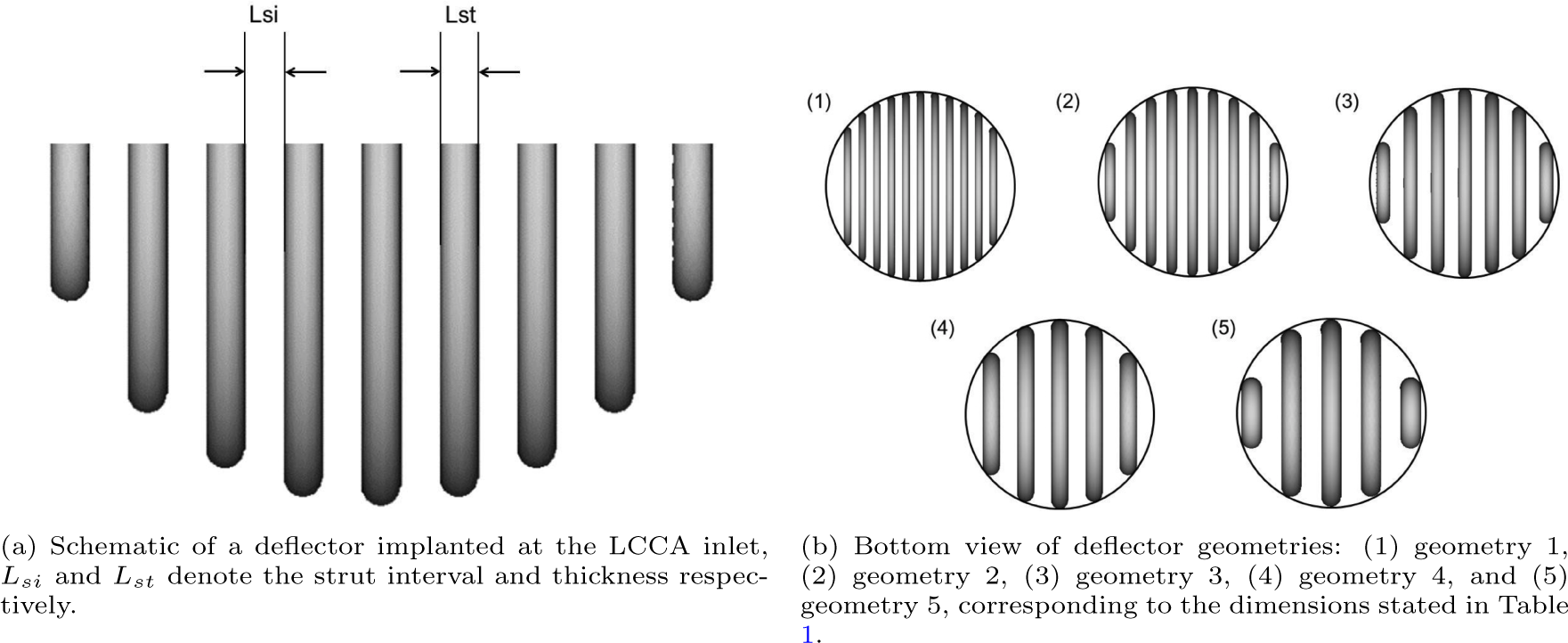
Schematics of device from different views.

**Table 1:** Dimensions for the design of the thrombus deflector for the different geometries generated.

| Deflector geometry | $L_{si}$ [mm] | $L_{st}$ [mm] |
| --- | --- | --- |
| Geometry 1 | 0.50 | 0.50 |
| Geometry 2 | 0.75 | 0.75 |
| Geometry 3 | 1.00 | 1.00 |
| Geometry 4 | 1.25 | 1.25 |
| Geometry 5 | 1.50 | 1.50 |

In order to merge the deflector device geometry to an aortic arch geometry, ANSA v22.1.0 (Beta Simulation Solutions, Lucerne, Switzerland) has been used. The deflector orientation angle with respect to the mean aortic flow was selected taking into account the results from [33]. In this study, the centerline of the aortic arch has been taken as the direction for the deflector positioning.

#### Lateral spacing

An important factor that has to be considered in the construction of the geometry and the implantation of the device is the lateral free space between the first struts of the deflector and the wall of the artery. As shown in Figure 6, the space between the lateral strut and the device annulus at the base of the LCCA varies among geometries, corresponding to 2.17, 1.76, 2.76, 1.9 and 1.76mm for the *L*_*si*_ = 0.5, 0.75, 1.0, 1.25, 1.5mm devices respectively. That is, all geometries have a bigger lateral spacing than their corresponding *L*_*si*_. The smaller lateral space of the *L*_*si*_ = 0.75mm design with respect to designs with *L*_*st*_ ∈ [0.5, 1.25] mm is consistent with the highest effectiveness in blocking particles from entering the LCCA as will be shown in Figure 15a. This is indicative of the critical impact of this geometric parameter in the design of the medical device.

**Figure 6:**
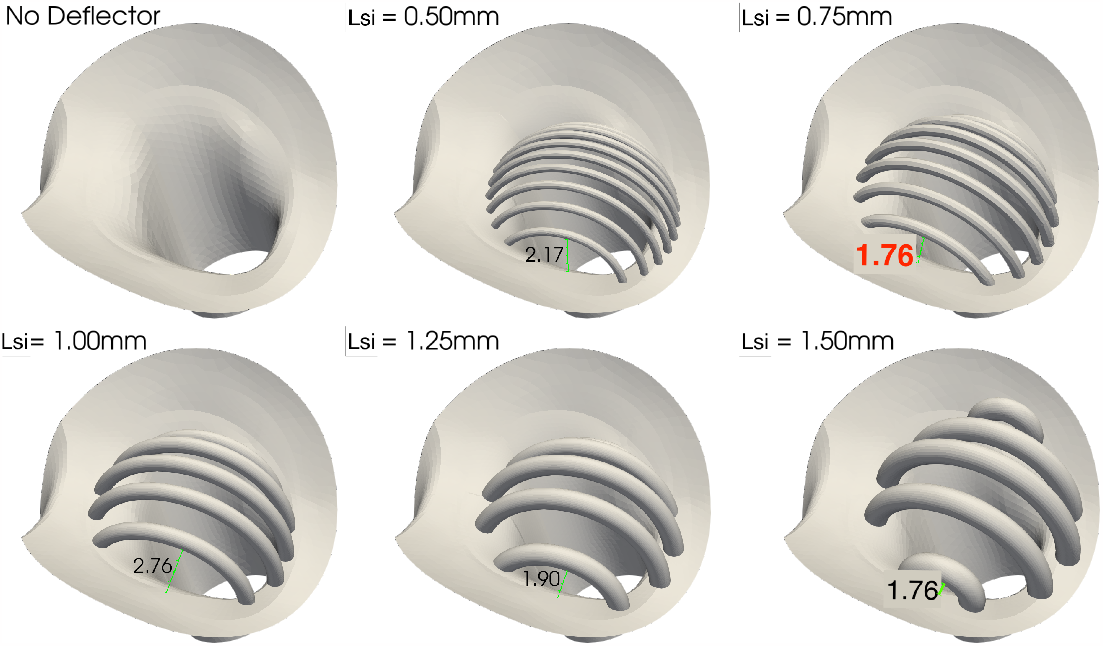
Deflector geometries of different strut thicknesses and inter-strut spacings. On top row from left to right: no deflector, *L*_*si*_ = 0.5 and 0.75mm bottom row: *L*_*si*_ = 1.0, 1.25 and 1.5mm. Spacings between lateral struts and annulus of base of LCCA are also displayed for each geometry. The *L*_*si*_ = 0.75 mm is of particular interest, since it is the most effective in blocking particles, with a lateral spacing of 1.76mm as highlighted in red.

Moreover, it is observed that some particles with larger diameter than the *L*_*si*_ of the device are able to enter the treated LCCA due to the above mentioned issue. Figure 7 shows a case where a large particle passes through the device and ends up in the LCCA. This extra lateral spacing explains the number of particles with diameter larger than *L*_*si*_ that get deposited in the LCCA output as well as the fluid jet that is also shown through this space in Figure 13 (top-left).

**Figure 7:**
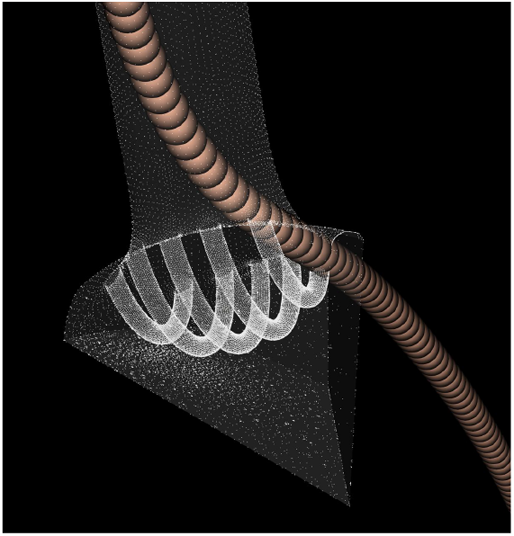
Particle with diameter larger than *L*_*si*_ passing through the device.

To overcome this deficiency, extra struts has been added in the new design employed in the second stage of the present work, oriented perpendicularly to the original struts.

#### Second stage

For the second stage of the present work, lateral struts were added to prevent particles from escaping through the side holes, the strut diameter was optimized and adapted to each of the three arteries.

Figure 8 shows the new struts added to the device to improve the particle filtering. It can be seen in this figure, that the strut thickness is not homogeneous, and instead thins out towards the ends (circled with red pointed lines). Note that this is an artefact of CAD model generation algorithm, in the creation of the 3D model, but is not a feature of the design or the patent involved. The thinning is an artefact of CAD model generation algorithm and in future versions of the design will be modified this morphology to maintain the section of the struts constant. Neither of the previous studies of this device had considered an electrical charge. In the second stage, electric charge is applied to additionally deflect small particles that could normally pass between the struts. The device would be covered with a negatively charged graphene oxide coating, with surface charges of −24000.0 statC cm^*−*2^, which corresponds to the most extreme electrical charges found in the literature of biomedical devices, as will be described later in section 2.3.2. [36].

**Figure 8:**
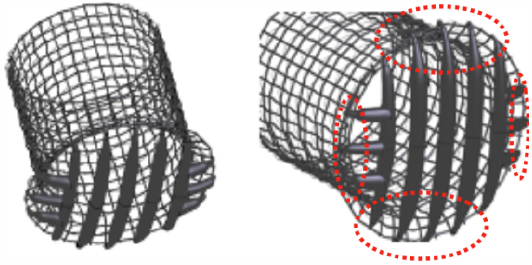
New deflector design. Lateral struts added are circled in red.

### 2.3 Physical model

#### 2.3.1 Blood flow model: computational fluid dynamics

##### Incompressible flow

Blood was modeled as a Newtonian incompressible viscous fluid. The Newtonian approximation is an acceptable assumption and not far from reality in a significant part of the circulatory system under normal conditions. This is especially true in large blood vessels where red blood cells are well below the characteristic sizes of the vessels [37]. The velocity and pressure fields in the aortic arch, ***u*** and *p* respectively, have thus been obtained by solving the incompressible Navier-Stokes momentum and continuity equations:

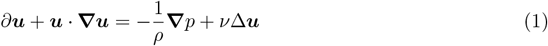

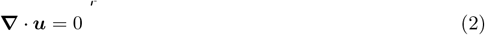

where 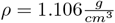 and 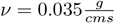 are the blood density and kinematic viscosity respectively. The corresponding boundary conditions are specified in Figure 2.

##### Boundary conditions for the flow: healthy and atrial fibrillation conditions

In order to set up the computational simulations, physical governing equations must be complemented by boundary conditions for both the fluid and particle dynamics as well as the electric field. For the fluid, the following boundary conditions are prescribed at the boundaries shown in Figure 2:

- Inlet: velocity Dirichlet boundary condition.
- Outlets: reduced-order (3-element Windkessel) model, imposed as a traction boundary condition, in which distal resistances and capacitances are modelled for a more accurate boundary conditions. In particular, the values for the present work are a 0 Pa for imposed pressure in the outlets, a resistive term equal to 100.0 and a scaling coefficient of 0.5.
- Arterial walls and deflector: no-slip Dirichlet boundary conditions.

It is noted that each one of the Windkessel boundary conditions were calibrated for the flow rates and pressures to fit the experimental results from [38] for the healthy patient case. The flowrate waveforms imposed at the inlet are representative of a healthy patient and an AF patient. In Figure 9 these two inflows used in the first stage are shown. The second stage modifies the AF inflow to adjust the physiology of a real patient in a more accurate way. Figure 9a the new inflow for AF is compared with the healthy patient. The healthy patient has a heart rate of 70 bpm and a cardiac output of 4.286 L min^*−*1^. In contrast, the AF patient has a higher heart rate, of 150 bpm, and a lower cardiac output, of 3.429 L min^*−*1^, a 20% decrease with respect to the healthy patient, as reported in [39].These flowrates were used for the second phase addapted to this specific geometry.

**Figure 9:**
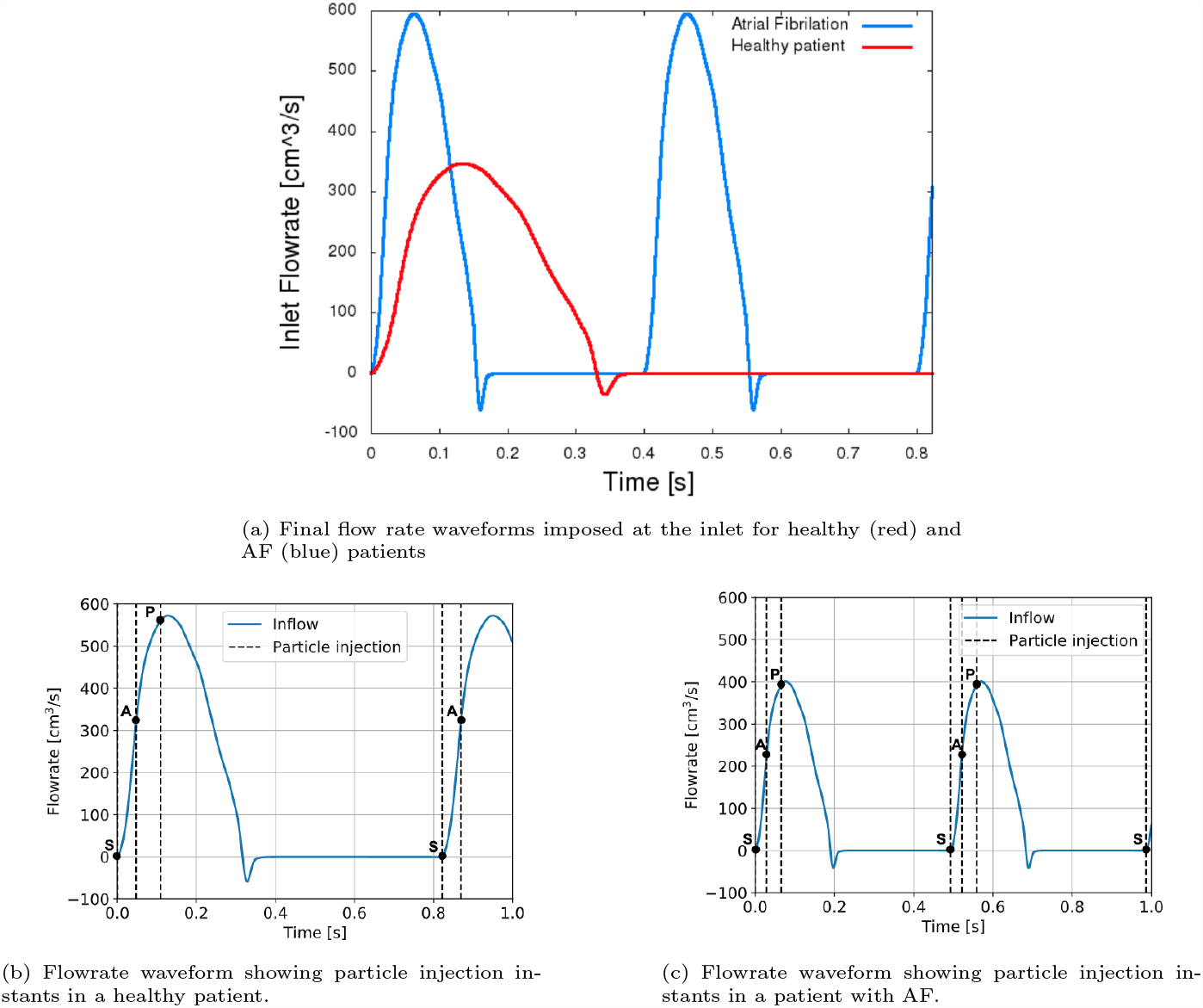
(a) Improved flowrate in the second stage of the work (above) and instants of particle injection, overlaid with inflow curve (below) (b) from a healthy patient and (c) from a patient with AF. Points in time S, A and P represent the beginning of systolic, accelerating, and peak stage of the cardiac cycle, respectively.

#### 2.3.2 Thrombus model: particle transport

Particle transport was simulated in a Lagrangian frame of reference, following each individual particle. The main assumptions of the model were:

- particles were sufficiently small and the suspension sufficiently diluted to neglect their effect on blood-flow: i.e. one way coupling;
- particles were spherical and did not interact with each other;
- particle rotation was negligible;
- thermophoretic forces were negligible; and
- the forces considered were drag *F*_*d*_, gravitational *F*_*g*_, buoyancy *F*_*b*_, and, for the second stage, electric force *F*_*e*_ is also considered.

Particles are injected assuming a circumferential distribution at a radial distance of 40% of the aortic radius [40], as observed in Figure 10. Clot injections are conducted at three different cardiac cycle moments assumed which correspond to a beginning of systolic (S), accelerating (A), and peak stage of the cardiac cycle (P) as in [40], illustrated in Figure 9.

**Figure 10:**
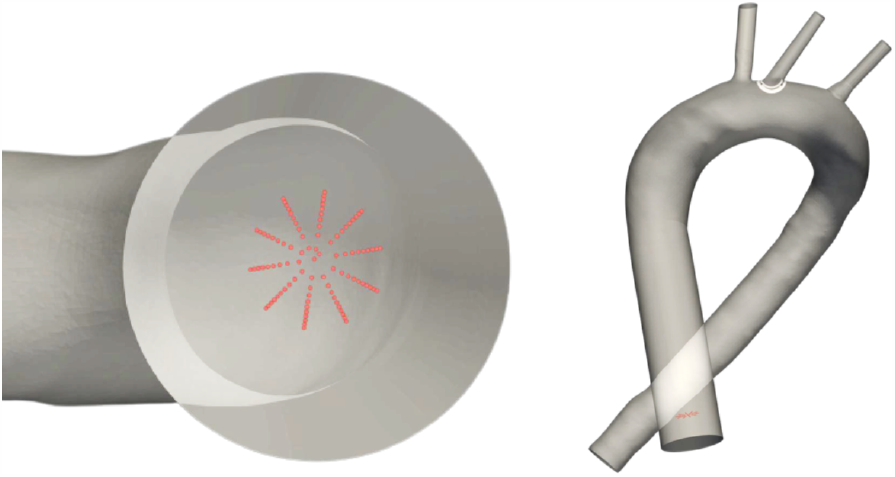
Zoomed view of the circumferential distribution (left) and vertical position at the inlet (right).

In order to properly define the deflection/blocking capacity of the device, different particle types, corresponding to different particle models defined by forces considered and the diameter and density properties for each type. These different particles were used as shown in Table 2.

**Table 2:** Particle types with their corresponding diameters (*dp*) and comparison with inter-strut spacing of deflector geometries (*L*_*si*_)

| Particle type | Diameter ($d_p$) | $d_p \geq L_{si}$ |
| --- | --- | --- |
| Type 1 | 10 $\mu\text{m}$ | None |
| Type 2 | 50 $\mu\text{m}$ | None |
| Type 3 | 100 $\mu\text{m}$ | None |
| Type 4 | 500 $\mu\text{m}$ | Geometry 1 |
| Type 5 | 1.00 mm | Geometries 1, 2 & 3 |
| Type 6 | 2.00 mm | All |
| Type 7 | - | N/A |

In particular all 7 were used in the first stage and only the first 5 (from 1st to 5th of the Table 2) in the second one. Force models are applied to the particles with types 1 to 6, which have a density of 1.08g cm^*−*3^ and different diameters, ranging from *d*_*p*_10 μm to 2mm. Particles of types 1 to 3 (*d*_*p*_ ∈ [10, 100]μm) are of importance for evaluating the efficacy of the device in avoiding micro-embolisms and associated SBI. Larger particles of types 4 to 6 (*d*_*p*_ ∈ [0.5, 2]mm) represent larger embolisms, which may produce cerebral strokes.

The trajectory of these particles can be obtained by using Newton’s Second Law, ***F*** = *m****a***, where the force ***F*** is the sum of the different forces exerted on the particle, *m* is the particle mass and ***a*** is the particle acceleration. In this case the forces applied on the particles are:

- **Gravity:** *F*_*g*_ = *m*_*p*_*g*, where *m*_*p*_ and *g* corresponds to particle mass and gravitational acceleration.
- **Buoyancy:** 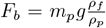, where *ρ*_*f*_ and *ρ*_*p*_ corresponds to the fluid and particle densities respectively.
- **Drag:** *F*_*d*_ the force acting in the opposite direction to the relative motion of an object moving with respect to a surrounding fluid. The equation for the drag force assumed the particle reached its terminal velocity and is given by:

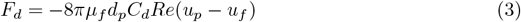

being *μ* the dynamic flow viscosity, *d*_*p*_ particle density and *Re* the particle Reynolds number, calculated from its relative velocity with the fluid: *Re* = |*u*_*p*_ − *u*_*f*_ |*d*_*p*_*/ν* with *u*_*p*_ and *u*_*f*_ corresponding to particle and fluid velocity. *C*_*d*_ is the drag coefficient given by Chen model described in [41]
- **Electric force:** *F*_*e*_ electrically charged deflector created an electric field which exerts a repulsive force through the electrical charged blood clots and described mathematically in the next section.

Particle transport was modelled by considering the above described forces, and solving Newton’s second law to obtain particle accelerations:

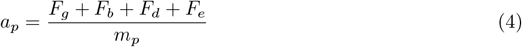

where *a*_*p*_ and *m*_*p*_ correspond to particle acceleration and particle mass.

For the case of type 7 particles, their trajectory is obtained by computing the velocity integral, meaning that these particles act like tracers, without considering any force acting on them and they move following the flow. All particles injected in the geometry are considered spheres with their diameters described in Table 2.

It is important to remark that *in vivo* studies are more likely to show larger clot diameters[42] than those specified in the table 2. However, those large clots were not considered in our analysis as they will simply not fit between the struts. Moreover, it is important to emphasise that as indicated in Table 2, for some deflector geometries, particles of types 4, 5 and 6 have a diameter larger than or equal to the inter-strut spacing (geometries 1, 2 & 3) and thus cannot enter the LCCA through this space.

##### Boundary conditions thrombus model

For the particles injected into the flow, the following boundary conditions are prescribed:

- **inlet & outlets:** absorbing boundary condition, which means particles passing through such surfaces.
- **arterial walls:** slip boundary condition, which allows tangential velocities and removes the normal one and
- **deflector:** elastic bounce boundary condition, which implies imposing normal velocity with opposite sense removing the tangential momentum.

##### Electrostatic field

The resulting electric field is obtained by solving the potential Poisson equation:

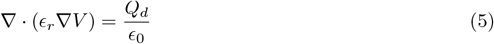

where *Q*_*d*_ is the total charge distributed superficially on the struts of the devices present in each configuration, *ϵ*_0_ and *ϵ*_*r*_ are the electrical permittivity for the vacuum and relative permittivity of blood. Finally, we use the relation between potential and electric field: *E* = −∇*V*. The electric field generated by the superposition of the three devices deployed in the three arteries is illustrated in Figure 11.

**Figure 11:**
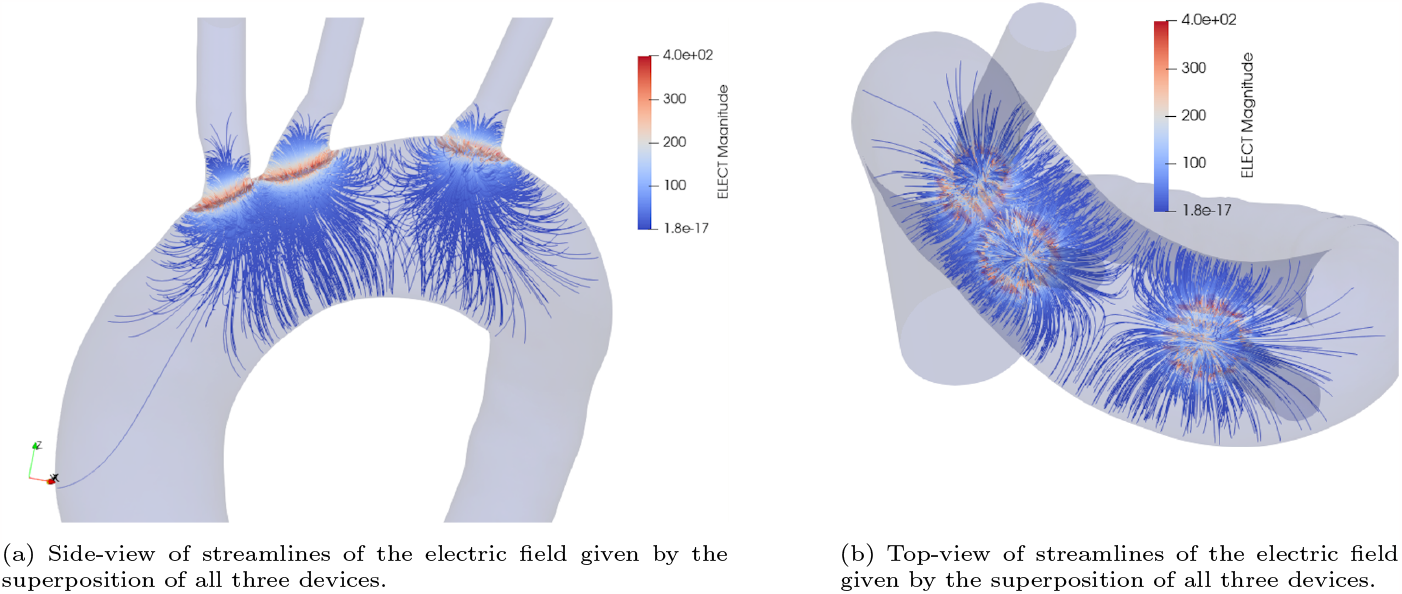
Streamlines of electric field generated by superposition of the 3 electrical charged devices placed in the 3 arteries

##### Boundary conditions electric potential

The boundary conditions of the Poisson equation are −1.3mV in the intima or internal aortic walls and free in the inlet and outlet surfaces.

#### 2.3.3 Electrical parameters of model

##### Blood flow model parameters

Blood viscosity, or the thickness of blood, is generally considered to be normal when it falls within a range of 3.5 to 5.5 cP. However, it is important to note that blood viscosity is not a constant value and can vary significantly depending on the specific conditions in which it is being measured. For example, the viscosity of blood can change based on the shear rate, or the speed at which the blood is flowing. At a shear rate of 0.1 s^*−*1^, the viscosity of the same blood sample may be as high as 60 cP. However, at a shear rate of 200 s^*−*1^, the viscosity would be much lower, at around 5 or 6 cP. This means that the viscosity of blood can differ in different parts of the circulatory system, such as the large arteries, veins, and microcirculation,where shear rates can range from a few s^*−*1^ to over 1000 s^*−*1^. Blood viscosity is influenced by several factors, including the concentration of red blood cells, the thickness of plasma, the ability of red blood cells to deform under flow, and their tendency to clump together [43], and therefore we considered blood visocisity value of 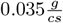

##### Thrombus model parameters

The charge of the simulated blood clots was determined following [44], where the authors study electrophoretic mobility of electrically charged particles. Electrophoresis involves the separation of charged clots under the influence of an electric field. In fact, this mechanism is the result of the combination of two different processes, intrinsic electrophoretic mobility and electroosmotic flow. The first one can be expressed as:

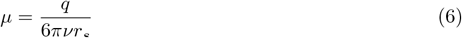

where *μ* is the electrophoretic mobility, q is the charge of the particle that we are interested in, *ν* the viscosity of the medium, and *r*_*s*_ the hydrodynamic radius. The electrophoretic mobility was taken as *μ* = −0.0336 cm^2^ statV^*−*1^s^*−*1^ = −1.12 μm s V^*−*1^ cm^*−*1^[45]. The densities assigned to blood clots were 1.08 g cm^*−*3^ [46]. By expressing the charge as a function of the remaining parameters in (6), electrical charges were obtained for each diameter, and summarized in Table 3.

**Table 3:** Particle diameters and their respective estimated electrical charges.

| Diameters [cm] | Charges [$\mu\text{F}$] |
| --- | --- |
| 0.001 | -11.0 |
| 0.005 | -55.0 |
| 0.01 | -111.0 |
| 0.05 | -554.0 |
| 0.1 | -1108.0 |
| 0.2 | -2215.0 |

##### Device model parameters

As described in Section 2.2, the surface charge on the graphene oxide in 0.001*MNaCl* was determined approximately as −75 mC m^*−*2^ in good agreement with the previous work [36] where the value was equal −80 mC m^*−*2^, at pH 6.5 extracted from a zeta potential measurement, albeit in NaCl. In CGS units, these two values correspond to −22, 484.43 statC cm^*−*2^ and −23, 983.39 statC cm^*−*2^. Based on these results, we have considered a surface charge of the device corresponding to −24, 000 statC cm^*−*2^. This value needs to be integrated for all the surfaces of the devices placed in the domain producing the total charge *Q*_*d*_ in (5), responsible for the electric field, which exerts the repulsive force on blood clots.

### 2.4 Quantities of interest

The following quantities of interest are used for both particle statistics and flow dynamics:

- The particle deflecting efficacy at the arterial outlets where the device is deployed, with and without considering electric charge. This metric provides a means of computing the efficacy of the device CEPD.
- The maximum ejected flowrate in the treated artery. This value can be used to determine the variation in flowrate ejected to the arteries with respect to the case without deflector. It thus enables computing what will be the effect on oxygen delivery to the cerebrovascular system.
- Volume-averaged metrics have also been computed in order to evaluate the risk of thrombus formation due to the presence of the medical device. The region of interest considered is the volume encapsulated by the deflector as shown in Figure 12. The quantities of interest evaluated are:
  - The average kinetic energy, associated to washout.
  - The average second invariant of the velocity gradient **V*u***, that is, 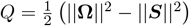, where ***S*** is the symmetric part of the velocity gradient known as the rate of strain, and **Ω** is the antisymmetric part known as the vorticity tensor. This invariant, is also used in the *Q*-criterion that quantifies the level of vorticity in the flow by defining vortices as volumes where *Q >* 0 (where the vorticity magnitude is greater than the magnitude of the rate of strain).
  - Δ*Q*:the drop in ejected flowrate at the arterial outlets where the device is deployed, relative to the untreated case.

**Figure 12:**
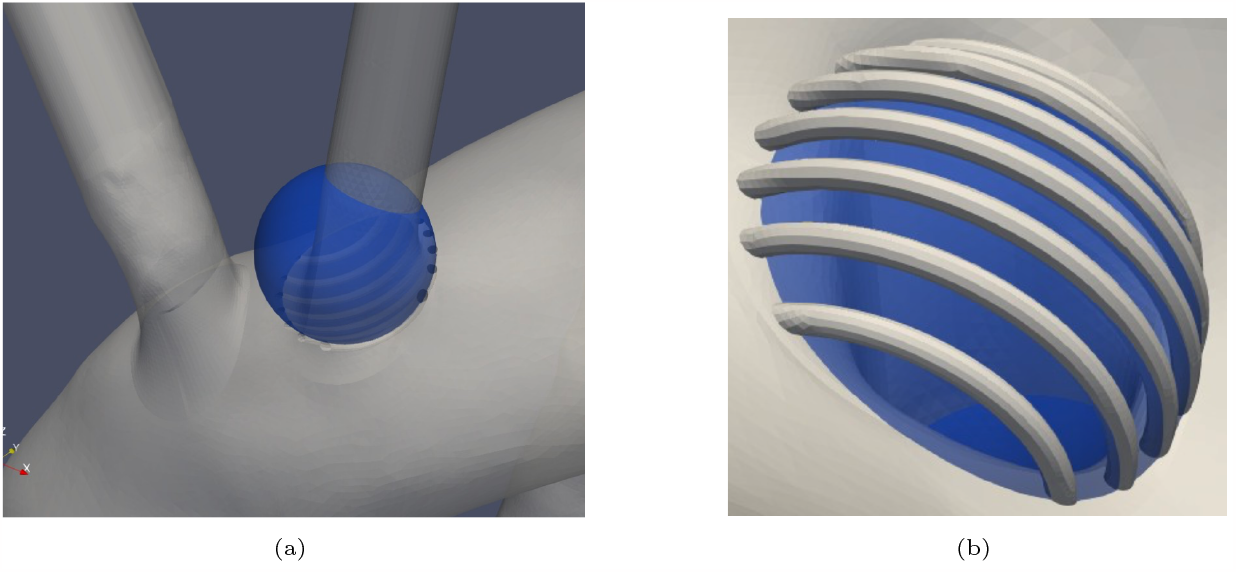
Region of interest where volumetric quantities of interest are integrated.

## 3 Results

### 3.1 Hydrodynamic quantities of interest

The first stage of the present work is focused on the analysis of the hydrodynamics effects of the deflector placed at the base of the arteries and how these hydrodynamics effects affect the trajectories of the potential clots travelling through the aortic arch. To that end, different configurations of the struts forming the presented device are analysed.

The presence of the deflector influences the haemodynamic flow locally. Figure 13 qualitatively depicts the fluid velocity field around the LCCA base where the deflector is deployed in this first study. Different instants of a single cycle are shown corresponding approximately to the peak stage (Figure 13a), the midterm of the deceleration phase (Figure 13b) and the plateau stage (Figure 13c). To quantitatively analyse the effect of the deflector on the flow, different hydrodynamic metrics were computed.

**Figure 13:**
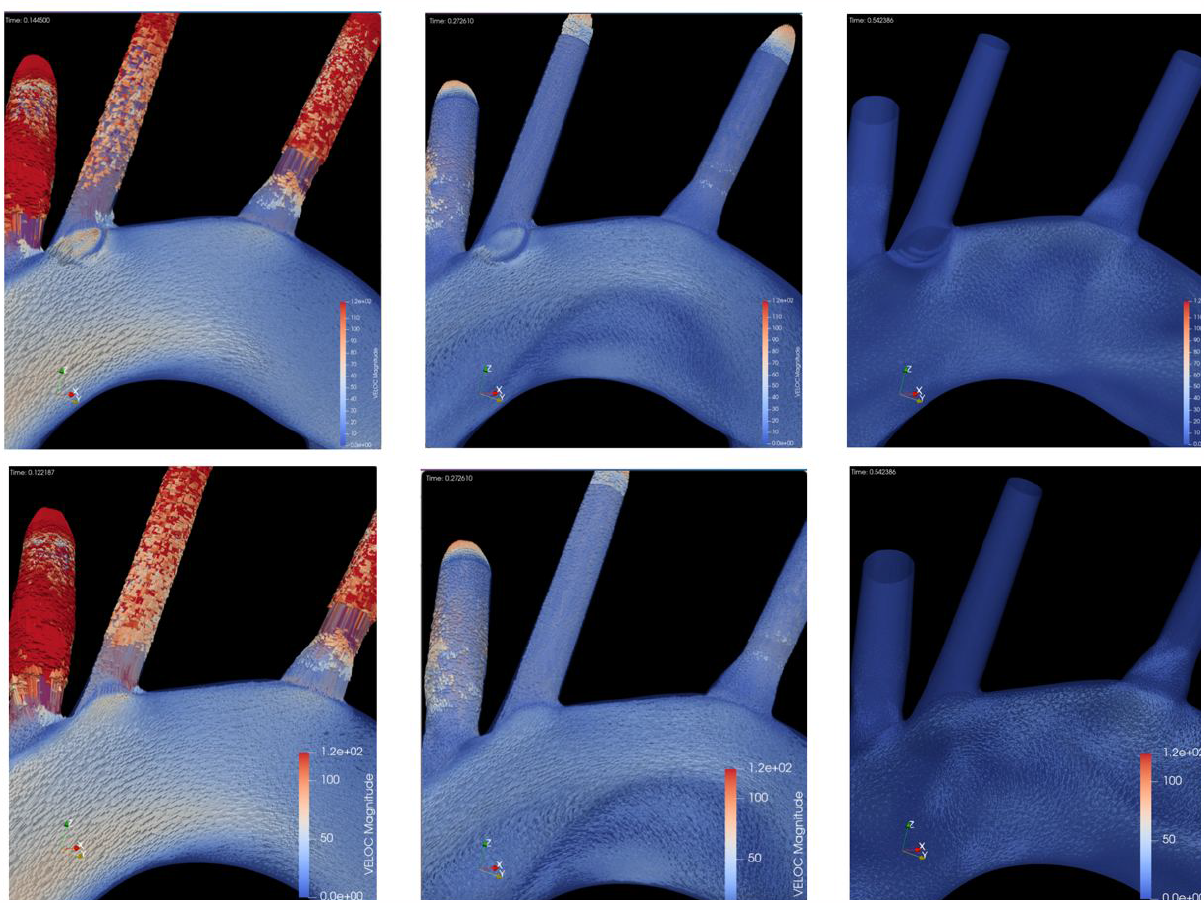
Velocity field around LCCA artery with deployed deflector (top) and without deflector device (bottom) for different instants of a single cycle, from left to right: (a) *t* = 0.1s, (b) *t* = 0.27s, and (c) *t* = 0.54s.

Considering the results of the second of the cited metrics, the maximun flowrate, we could observe that the mean value all the configurations is 68,91 with a standard deviation of 1.019. So no significant reduction in flowrate is obtained in either case for the different configurations represented in Table 1.

The kinetic energy quantity of interest evaluated in the region of interest considered in Figure 12 is reduced comparing the non-device configuration for all the analyzed configuration, obtaining a maximum value of 20% for the Geometry-4 case of Table 1. Although the results indicates that the presence of the deflector reduces the velocity downstream of the deflector, the Geometry-2 configuration is the one that mantain higher kinetic energy with a reduction of less that 10 %.

In order to quantify the degree of vortices generated by the deflector, the Q-invariant (second invariant of velocity gradient) is computed. Based on the results obtained, again we see that the deflector presenting a lowest *Q* value (*i*.*e*., presenting similar results to the case with no deflector implanted) is the one with dimensions *L*_*si*_ = 0.75 mm and *L*_*st*_ = 0.75 mm, being the design which produces less vortices in the evaluated region.

### 3.2 Particle deflecting efficacy

The subsequent results comes from the two stages of the presented work:

- For the first stage, with no electrical forces and as it has been described in Section 4.3 and illustrated in 5b, the analysed particle deflecting efficacy is focused in the behaviour of the geometric design parameters: the distance between deflector struts or strut interval (*L*_*si*_) and the strut thickness (*L*_*st*_). The deflector is only located at one artery, LCCA, and 5 different prototypes based on mentioned parameters are analysed.
- The second stage of the work, now with electrical forces computed, covers the effect of multiple devices deployed at the same time. In particular five different device deployment configurations shown in Figure 14 are presented, which corresponds from non-deflector (14-a) case to deflector located in all 3 arteries (14-e), considering also the singles cases, this means deploying a single deflector at the base of the BCT only (14-b), at the base of the LCCA only (14-c) and at the base of the LSA only (14-d). All of these also consider the effect of the electric field produced when the device is recovered by an electrically material. For each case, a total of 6 million particles were injected into the domain during the full length of each simulation. This ensemble of particles is equally distributed among 10 different particle types, that is, injecting 150,000 particles of each type during the full length of each simulation. The 10 particle types are a result of the combination of 5 particle sizes and 2 electrical charge conditions. The particle injections are distributed in time, with 3 injections per cardiac cycle. In each injection, 10,000 particles of each type were introduced into the inlet, homogeneously distributed in space. The 5 different particle diameters considered here are the ones presented in Table 2 except the last two, that has been avoided now due to the dimensions of the chosen configuration in the previous stage.

**Figure 14:**
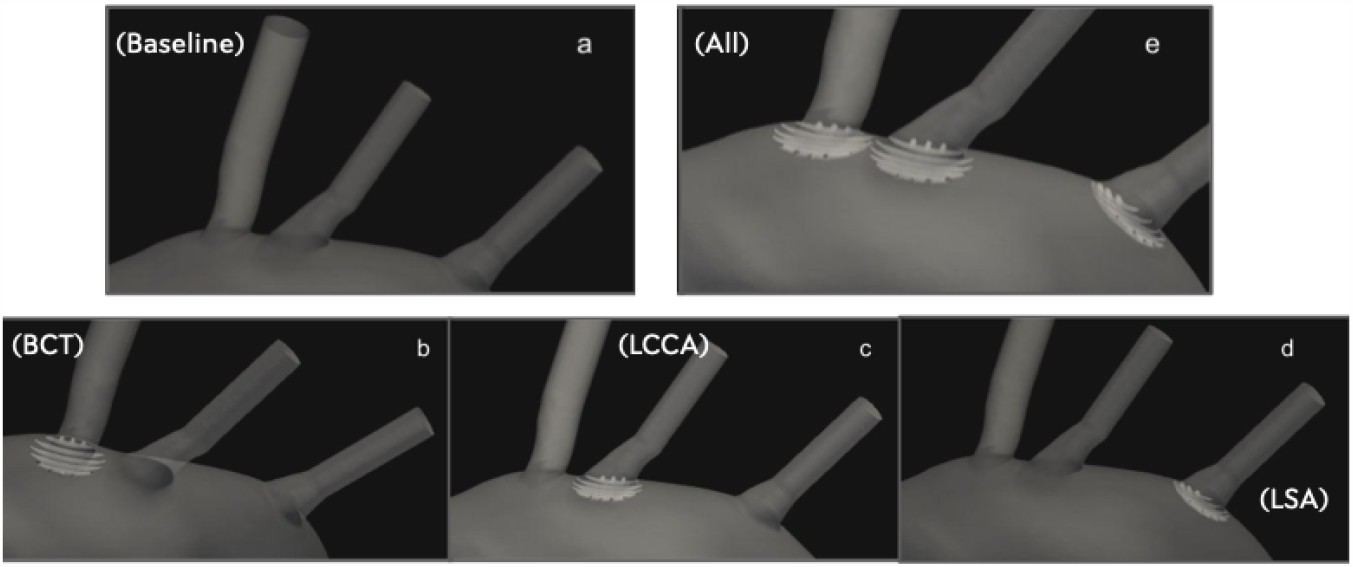
Five different device deployment configurations: (a) No deflector. (b) Only in BCT. (c) Only in LCCA. (d) Only in LSA. (e) In 3 arteries

#### 3.2.1 Healthy patient

##### Comparison between all different morphologies over LCCA

Figure 15 shows results for the particles effectively passing through the deflector device (in between struts) deployed at the base of the LCCA. Particle counts are normalised by the total number of particles reaching the LCCA outflow in the no-deflector case, obtaining our quantity of interest called *ϵ*. It can be seen that particle counts there are only reduced for particles above or equal to 0.5 mm for the healthy patient case (Figure 15a). These sizes correspond to the minimum inter-strut spacings of the analysed designs. As should occur, particles above the inter-strut spacing are 100% blocked, while when slightly below the inter-strut spacing (*e*.*g*., see Figure 15a: *d*_*p*_0.5 mm, *L*_*si*_ = 0.75 mm where particle deposition is reduced by 38%), a moderate reduction is observed. For particles well below the inter-strut spacing (*d*_*p*_ ≪ *L*_*st*_) no significant reduction is observed. In fact, for some of these particle-strut combinations an increased number of particles are detected in the LCCA. It is remarkable that reductions in particles passing through the device are observed for particles with diameters below *L*_*si*_ in the *L*_*si*_ = 0.75 mm in the healthy case (Figure 15a).

**Figure 15:**
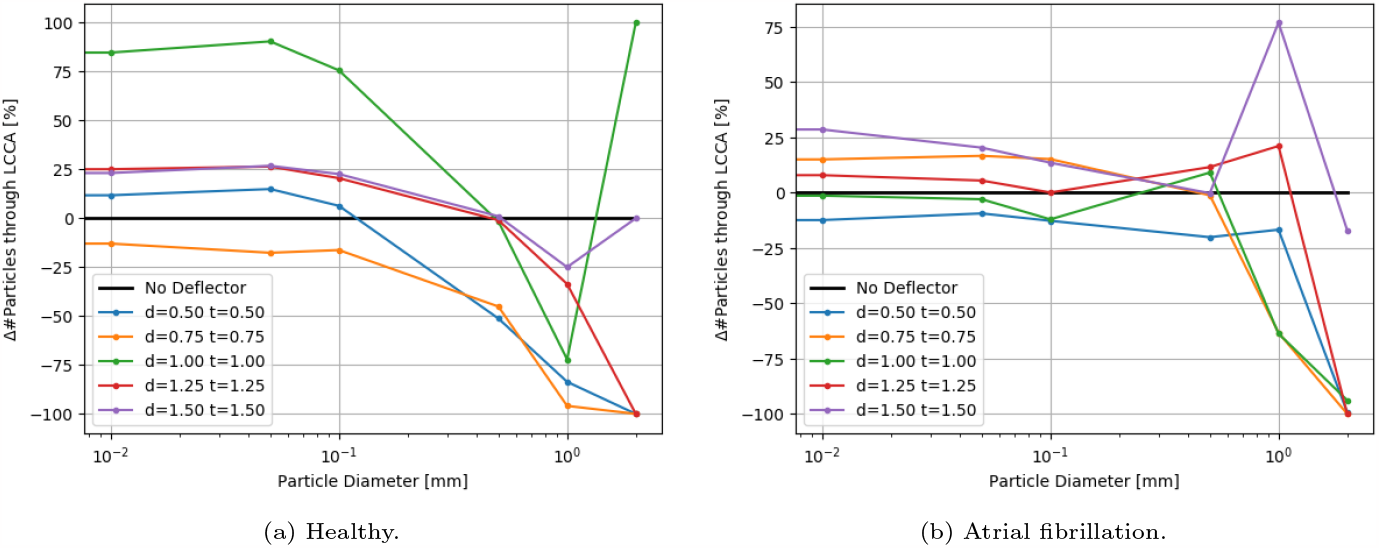
Variation in number of particles entering the LCCA for increasing strut thicknesses for the (a) healthy, and AF patient. Particle depositions are normalised by the no-deflector case *L*_*si*_ = 0mm. Inter-strut spacings are equal to the strut thickness in each case, as can be seen in Table 1.

##### Comparison between electric field repulsion and no-charged device for the optimum configuration of first stage

In this second stage of the work, as it is described below, 10 simulations are carried out. In each of one, for each size of the particles, 2 electrical charge conditions were considered: neutral and electrically charged. The thrombus deflecting performance of the electrically charged device was evaluated with a flow rate waveform representative of a healthy patient (see Figure 9a). This waveform corresponds to a heart rate of 70 bpm and a cardiac output of 4.286 L min^*−*1^. As a baseline, the simulation was first run without deploying the deflector (Figure 14a), recording the number of particles of each type exiting each arterial outlet of the domain. Then, the device was deployed individually in each one of the 3 aortic arch arteries, repeating the process (Figures 14b, c, d), with particle filtering efficacies shown in Table 4 4a. Finally, the device was deployed in all 3 aortic arch arteries simultaneously (Figure 14e), with the particle filtering efficacies shown in Table 5 4b. The absolute number of particles of each type exiting each artery were recorded and are given in Table 6a of the appendix.

**Table 4:**
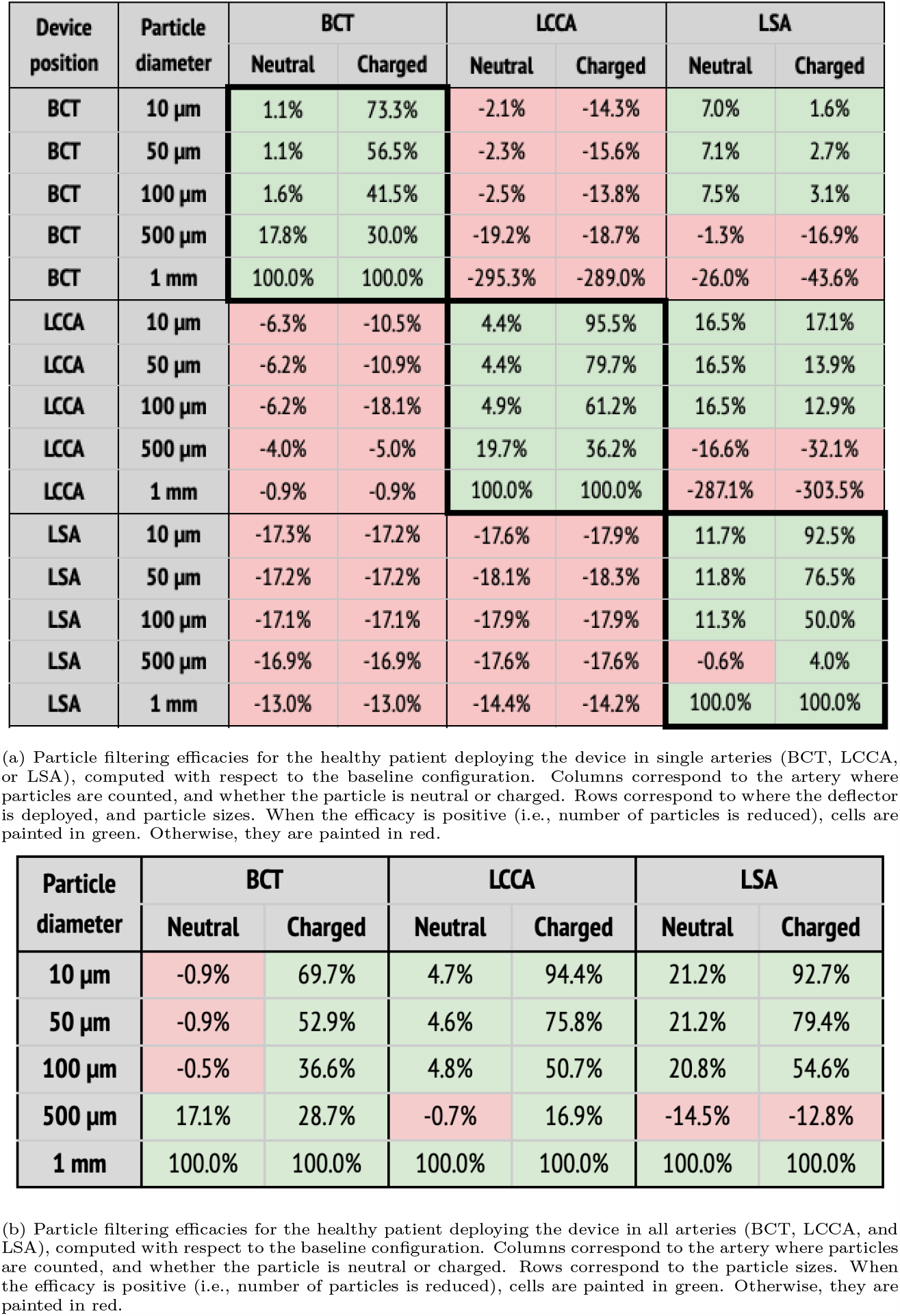
Particle filtering efficacies for healthy patients

**Table 5:** Particle filtering efficacies for the AF patient

| Device position | Particle diameter | BCT |  | LCCA |  | LSA |  |
| --- | --- | --- | --- | --- | --- | --- | --- |
|  |  | Neutral | Charged | Neutral | Charged | Neutral | Charged |
| BCT | 10 $\mu\text{m}$ | 10.5% | 63.1% | -18.1% | -26.6% | -15.3% | -18.6% |
| BCT | 50 $\mu\text{m}$ | 10.6% | 46.3% | -18.3% | -27.6% | -15.0% | -18.0% |
| BCT | 100 $\mu\text{m}$ | 10.9% | 35.9% | -18.8% | -27.7% | -14.3% | -17.2% |
| BCT | 500 $\mu\text{m}$ | 15.9% | 21.2% | -24.4% | -24.1% | 13.5% | 7.2% |
| BCT | 1 mm | 100.0% | 100.0% | -328.5% | -327.2% | 13.0% | 4.0% |
| LCCA | 10 $\mu\text{m}$ | -9.9% | -13.8% | 4.0% | 76.5% | 18.9% | 16.8% |
| LCCA | 50 $\mu\text{m}$ | -10.1% | -20.4% | 4.8% | 55.3% | 18.7% | 15.2% |
| LCCA | 100 $\mu\text{m}$ | -10.8% | -26.5% | 8.0% | 45.2% | 16.9% | 14.2% |
| LCCA | 500 $\mu\text{m}$ | -2.2% | -3.6% | 42.0% | 46.3% | -18.4% | -20.7% |
| LCCA | 1 mm | -0.2% | -0.8% | 100.0% | 100.0% | -169.4% | -174.2% |
| LSA | 10 $\mu\text{m}$ | -9.7% | -9.7% | -6.9% | -6.8% | 16.2% | 79.8% |
| LSA | 50 $\mu\text{m}$ | -10.0% | -10.0% | -6.3% | -6.2% | 17.1% | 60.3% |
| LSA | 100 $\mu\text{m}$ | -9.4% | -10.2% | -5.6% | -5.7% | 18.0% | 38.6% |
| LSA | 500 $\mu\text{m}$ | -9.4% | -9.4% | 11.1% | 11.2% | 22.8% | 25.9% |
| LSA | 1 mm | -5.8% | -5.8% | 7.0% | 6.9% | 100.0% | 100.0% |

| Particle diameter | BCT |  | LCCA |  | LSA |  |
| --- | --- | --- | --- | --- | --- | --- |
|  | Neutral | Charged | Neutral | Charged | Neutral | Charged |
| 10 $\mu\text{m}$ | -6.0% | 54.4% | -4.2% | 73.5% | 11.7% | 80.8% |
| 50 $\mu\text{m}$ | -5.0% | 31.7% | -4.4% | 43.6% | 12.3% | 61.8% |
| 100 $\mu\text{m}$ | -5.1% | 10.0% | -2.1% | 25.9% | 12.7% | 41.1% |
| 500 $\mu\text{m}$ | 17.3% | 20.1% | 13.7% | 20.1% | 2.9% | 4.9% |
| 1 mm | 100.0% | 100.0% | 100.0% | 100.0% | 100.0% | 100.0% |

(b) Particle filtering efficacies for the AF patient deploying the device in all arteries (BCT, LCCA, and LSA), computed with respect to the baseline configuration. Columns correspond to the artery where particles are counted, and whether the particle is neutral or charged. Rows correspond to the particle sizes. When the efficacy is positive (i.e., number of particles is reduced), cells are painted in green. Otherwise, they are painted in red.

When the deflector is implanted in a single artery, it effectively reduces the number of particles there, producing increases in the untreated arteries (Table 4a). Moreover, the filtering effect is improved when the electrical charge is considered. When deploying the device in all arteries, the number of particles are also reduced in all terminals, on average by 62.6%, but it is necessary to consider the electrical charge in order to produce the desired effect (Table 4b).

#### 3.2.2 Atrial fibrillation patient

##### Comparison between all different morphologies over LCCA

In this case, as we can see in Figure 15b, particle counts there are only reduced for particles above or equal to 1.0mm for the AF case. The reductions in particles passing through the device are observed in Figure 15b with diameters below *L*_*si*_ = 0.5mm in this AF case.

##### Comparison between electric field repulsion and no-charged device for the optimum configuration of first stage

In a second step, a flow rate waveform representative of an AF patient was imposed at the inlet. This waveform corresponds to a heart rate of 150 bpm and a cardiac output of 3.429 L min^*−*1^, which corresponds to a 20% decrease with respect to the healthy patient [39]. As for the previous section, the five deployment setups shown in Figure 14 were considered: baseline (no deflector), deploying the device only in the BCT, only in the LCCA, only in the LSA, and in all three arteries simultaneously. The particle filtering efficacies for the deflector deployed in single arteries are shown in Table 5a, while Table 5b gives the efficacies for the deflector deployed in all arteries at once. The absolute particle counts in each artery are shown in Table 7a of the appendix.

As observed for the healthy patient, in the diseased patient deploying the deflector in single arteries filters particles, even without considering electrical charge (see Table 5a). Nonetheless, only significant particle filtering efficacies (between 21.2% and 100%) are observed when the electrical charge is activated. As seen for the healthy patient, this has the collateral effect of increasing the particle counts in the untreated arteries. On the other hand, when deploying the deflectors in all arteries at once, the electrical charge is necessarily required to filter out particles, with an average efficacy of 51.2% (see Table 5b).

## 4 Discussion

### 4.1 Device efficacy in the healthy patient

In this section the device performance is analyzed for the healthy flow rate conditions, in the baseline (i.e., no deflector), deploying a single deflector on each one of the aortic arch arteries separately, and finally deploying the deflectors in all arteries at once.

#### 4.1.1 Baseline: no deflector

In the baseline configuration, that is without any deflector deployed (Figure 7a), it can be observed in Figure 8 that for the smallest particles (10 μm to 100 μm), the number of particles exiting each outlet is inversely related to the proximal distance to the aortic root (in increasing order: BCT, LCCA, LSA, and DAO). This tendency is reversed for the largest particles (500 μm to 1 mm), for which the number of particles at each outlet increases with the proximal distance instead. This observation can be explained by the fact that as the particles get larger, their inertia dominates over the flow advection, resisting to curve their trajectory, and thus entering the first outlet that they encounter along their straight path. On the other end, smaller particles are dominated by the flow advection, and thus are more evenly distributed between the outlets, with most particles exiting the widest outlet, the DAO.

#### 4.1.2 Deflector deployed in a single artery

Following the baseline case, the deflector was deployed individually in each aortic arch artery (BCT, LCCA, or LSA), running separate simulations for each case (Figures 14b, c, d respectively). These setups are intended to isolate the effect of the deflector on each artery and to quantify the collateral effect on the complementary arteries. Here, the complementary arteries refers to the arteries left untreated (e.g., BCT and LSA), when the deflector is deployed in a given artery (e.g., LCCA). A significant reduction in the particle counts is observed for the treated arteries (thick borders in Table 4a), from the neutral particles to their charged counterparts. Five main observations are noted with respect to this effect:

1. the neutral particles are filtered with an efficacy of 26% on average in the artery where the deflector is deployed,
2. this average efficacy increases to 66.5% when the particles are charged,
3. the deflection efficacy of the electrical charge diminishes for larger particle sizes,
4. the number of particle counts of arteries distal to the artery where the deflector is deployed increase by 38.8% on average with respect to the baseline case (no deflector), and
5. they increase to 46.6% when the particles are electrically charged.

Regarding the third point, the deflection of particles is most effective for the smallest particles (10 μm), which reduce their counts by 73.3%, 95.5%, and 92.5% at the BCT, LCCA, and LSA, when deploying the deflector in each one of these arteries respectively. Note that this effect is observed for all particle sizes, except for the largest (1 mm), which are directly filtered because they can’t fit in between the struts, and exit through the DAO instead. Regarding the second and third observations listed above, it can be seen in Table 4a that deploying the deflector in a given artery increases the number of particles in the free distal arteries by 38.8% on average, while only increasing the particle counts by 12.7% in the proximal arteries. This effect is enhanced when the particles are electrically charged, further increasing particle counts by 46.4% in the distal arteries, and 14.2% in the proximal arteries. The fact that deploying the deflector in a single artery may have negative consequences in either of the remaining aortic arch arteries, motivates the deployment of the device in all three arteries, as described in the following section.

#### 4.1.3 Deflectors deployed in all three arteries

Three deflectors were deployed in all arteries (BCT, LCCA, and LSA) simultaneously, imposing the healthy patient flow rate curve at the inlet. This setup is intended to fully protect the cerebrovascular circulatory system from cardioembolisms coming from the aortic root. Table 4b shows the corresponding particle counts in each one of the arterial outlets, which are plotted in Figures 16b,c,d for the BCT, LCCA, and LSA respectively. Electrically charging the device struts reduces the number of particles entering the aortic arch arteries in all cases, except for the largest particles (1 mm), which are blocked with or without charge, since they can’t physically fit between the struts (the inter-strut spacing is 0.75mm). On the other end of the size spectrum, as for the treated arteries in section corresponding on the single artery study, the electrical deflection mechanism is strongest for the smallest particles (10 μm), and is diminished as the particles get larger and inertia increasingly dominates over electrostatic forces. Interestingly, except for the 500 μm particles, the device has a two-fold deflecting/filtering mechanism: while the smallest particles are deflected by electrostatic repulsion, the largest ones are mechanically filtered since their diameter is similar or larger than the inter-strut spacing. In consequence, the device effectively avoids a wide range of particle sizes from entering the aortic arch arteries, protecting the cerebrovascular system. For the 500 μm particles, the effect of the device located in all three arteries leads to a unexpected behavior in the LSA, where it actually increases the number of particles entering the artery (Figure 16d). This issue must be addressed in further detail in future iterations. It is nonethelesss worth mentioning that the effect of the electric field attenuates the problem presented in this case.

**Figure 16:**
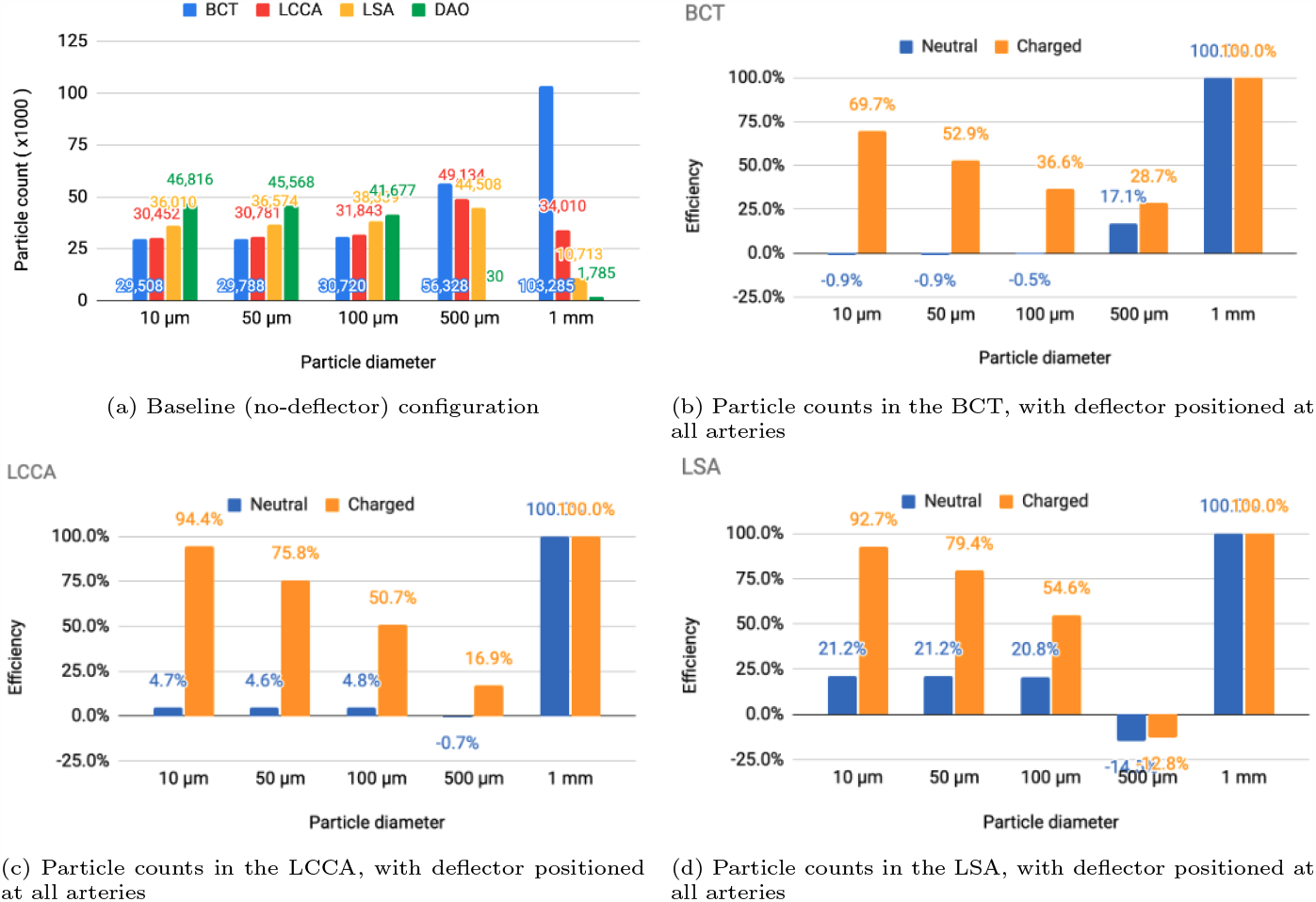
Particle counts of the healthy patient.

### 4.2 Device efficacy in atrial fibrillation patient

In this section the device efficacy is analyzed for the AF flow rate conditions, in the baseline (no deflector, Figure 14a), deploying a single deflector on each one of the aortic arch arteries separately (Figures 14b, c, d), and finally deploying the deflectors in all arteries at once (Figure 14e).

#### 4.2.1 Baseline: no deflector

As for previous section, the baseline configuration is examined for the AF case, that is without any deflector deployed (Figure 14a). As for the healthy patient, Figure 17a shows that the particle counts for the smallest particles (10 to 100 μm) are inversely related to the proximal distance to the aortic root. For the larger particles (500 μm to 1 mm), the inverse tendency which was observed for the healthy patient, is also observed in the AF patient in Figure 17a. The fact that the behaviour is conserved, indicates that the tendencies in final particle positions are maintained from the healthy to the diseased patient, in a statistical sense.

**Figure 17:**
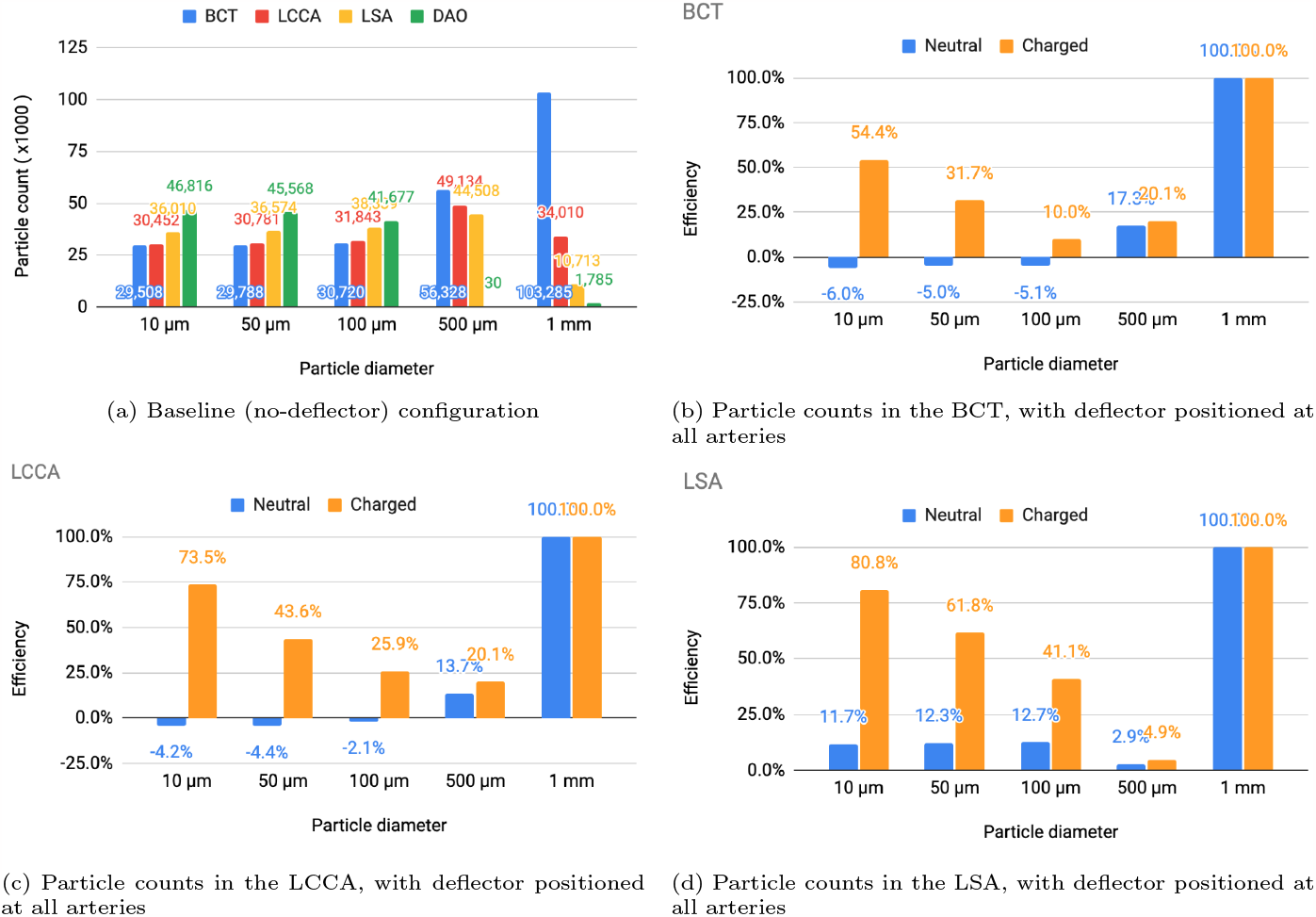
Particle counts of the healthy patient.

**Figure 18:**
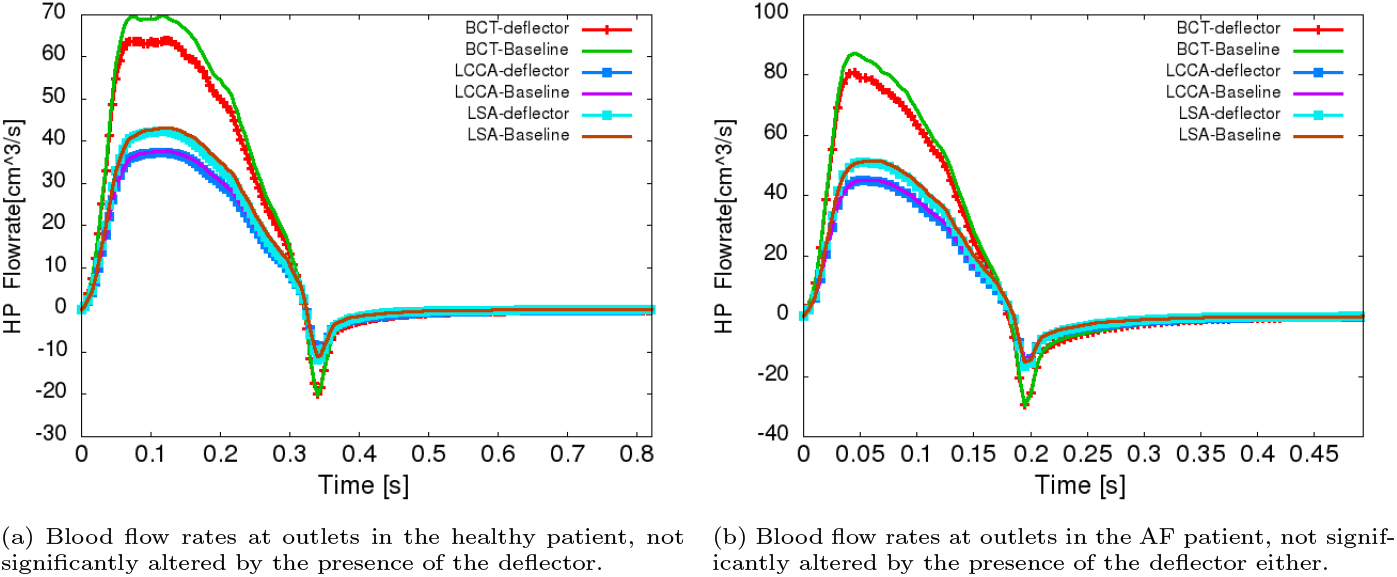
Safety flowrates. Reduction of the blood flow rate not significantly into the aortic arch arteries

#### 4.2.2 Deflector deployed in a single artery

Analogously to previous section, the deflector was deployed individually in each aortic arch artery (BCT, LCCA, or LSA), running separate simulations for each case (Figures 14b, c, d respectively), prescribing the AF flow rate at the inlet. The observations noted in the healthy patient are still observed for the AF patient, with similar particle count differentials when implanting the deflector, and when considering electrical charge:

1. the neutral particles are filtered with an efficacy of 32.0% on average in the artery where the deflector is deployed (26.0% for the healthy patient),
2. this average efficacy increases to 59.6% when the particles are charged (66.5% for the healthy patient),
3. the deflection efficacy of the electrical charge diminishes for larger particle sizes,
4. the number of particle counts of arteries distal to the artery where the deflector is deployed increases by 37.3% on average with respect to the baseline case (38.8% for the healthy patient), and
5. they increase to 41.6% when the particles are electrically charged (46.6% for the healthy patient).

In summary, as for the healthy patient, in the AF patient, the deflector effectively reduces the particle counts in the treated arteries, while producing a slight increase of the particles entering distal arteries left open.

#### 4.2.3 Deflectors deployed in all three arteries

When deploying the deflector in all three arteries in the AF conditions, similar results are observed to those seen in the analogous case of healthy patient. Table 5b shows the corresponding particle counts in each one of the arterial outlets, which are plotted in Figures 17b,c,d for the BCT, LCCA, and LSA respectively.This is, the deflector effectively reduces the number of neutral particles entering the treated aortic arch arteries, by 22.9% on average. When the electrical charge is incorporated, the filtering efficacy further increases, to 51.2%. As in previous sections, the largest particles (μm) are blocked with or without charge, since they can’t physically fit between the struts (the inter-strut spacing is 0.75mm). Therefore, once again the two-fold deflecting/filtering mechanism of the device is evidenced: while the smallest particles are deflected by electrostatic repulsion, the largest ones are mechanically filtered since their diameter is similar or larger than the inter-strut spacing.

### 4.3 Device safety

One of the main necessary conditions to assure the safety of the device, is that the presence of the deflectors does not reduce the blood flow rate significantly into the aortic arch arteries, necessary to provide the brain with oxygenated blood. Figures 16 and 17 overlap the flowrate curves at these arteries, in the baseline case (without deflector) and in the case where the three devices are deployed simultaneously, both for the healthy and AF patients. These correspond to the worst case scenarios for reduction of the flow, and therefore are sufficient to assure the device safety when the device is deployed in a single or two arteries. From these curves, it is observed that the device does not alter the flow rates significantly in any case. The maximum reduction is found in the BCT artery, with a 8.63% reduction for AF patients and 8.64% for healthy ones, with respect to the baseline case. In the LCCA artery the reduction is equal to 1.61% and 1.59% for AF and healthy patients while for LSA there is 4.03% and 3.97% less flow for the AF and healthy patients respectively when the device is deployed. In summary, the model predicts that the device reduces the ejected flowrate to the aortic arch arteries by significantly less than 20%, which is the maximum tolerated physiological reduction. Therefore, the device complies with the necessary condition set at the beginning of this report, which is required to assure a healthy cerebrovascular function of the treated patient.

### 4.4 Clogging of the device

Particles blocked by the mere fact that their diameter is larger than the inter-strut spacing may cause concern regarding clogging of the device, as occurs with conventional CEPD. The current numerical model cannot evaluate this aspect since deformability of the particles is not modelled. Nonetheless, it is somewhat reassuring that in contrast to conventional CEPD, the current device has a positive convexity which slightly bulges into the main aortic domain. This geometric attribute combined with the high velocities in the aorta result lower the possibility of particles getting stuck in between struts.

This effect is also addressed and improved introducing the repulsive electrical force due to the inverse distance proportional factor presented in this force.

## 5 Limitations

One of the limitations in the mathematical model considered is to assume clots as point-like particles for the fluid-particle interaction. As it was described in 2.3.2, the tracking is obtained by integrating the second Newton’s law, so, mass affects in the effect of the forces but not shape. Moreover, the fluid-particle coupling is one-way, that is, particles do not influence the fluid flow. These two approximations are not accurate for particles well above the Kolmogorov scale of the flow. Therefore, errors introduced by the point-like approximation can be seen to increase with particle size. For the collisions of particles with the domain walls, particles were modelled as perfect spheres, and slip boundary conditions were imposed. The rigidity and shape of spherical particles may introduce further errors in the model when compared to blood clots which are deformable and amorphous. The slip boundary conditions have shown more realistic results than considering boundary conditions, but still do not fully reproduce the interaction of blood clots with the aortic lumen, which could be improved.

In addition to limitations in the mathematical model, the physical parameters used in the present work are based on the scarce references found in literature, of limited credibility. Therefore, to assure a higher credibility of the results, these parameters require further verification and validation with experimental data. Some examples of these parameters that are complex to determine, are the permittivity of the blood, the electric charge and diameters of the clots, and the charge of the strut graphene oxide coating. Moreover, it is necessary to clearly determine the physiological limits for the device electrostatic charge, strong enough to produce a significant effect on particles, but not so strong so as to onset hemolysis or cause negative physiological consequences.

Regarding the geometrical model of the device, due to an artefact introduced in the CAD generation, the side struts were modelled with a heterogeneous thickness, thinning out towards the ends. This implies that free space greater than 1 milimeter is found in the device. This feature is hypothesized to have produced the spurious effect of actually increasing the number of 500 μm particles entering the treated LSA in the healthy case. This is, nonetheless, a model that will be evaluated and improved in further phases. The present study is a proof of concept that demonstrates the potential of the proposed device and the methodology to be optimized, but, given the novelty and risk of the proposed device, it is imperative to carry out a thorough experimental validation.

## 6 Conclusions

The present study carry out a computational analysis of a thrombus diverting and filtering device, designed as a solution to stroke and SBIs, critical health issues which are becoming more frequent given aging populations, increasing prevalence of AF, and devices being deployed in increasingly younger patients. The work has been carried out in two main stages. The first one analyzed the effect of the thickness and shape of the strut design on the device performance. The purely hydrodynamic effect of the device was analyzed using a CFD and particle transport model. The device was placed at the root of the LCCA and the optimal strut thickness was identified by analyzing the trajectories of particles suspended in the flow. The analysis concluded a low efficacy for the deflection of thrombi and identified a deficiency in the initial design which was filtering particles within the lateral struts. To overcome this deficiency, extra struts were added in the new design used in the second stage of the present study, oriented perpendicularly to the original struts.

In the second phase, given the electronegative charge of blood clots, a negative electrostatic charge is imposed on the deflector struts in the device design, in order to repel thrombi. The computational model employed for this analysis combines computational fluid dynamics and electrostatics. The numerical analysis results showed that, while maintaining the patient models’ blood flow at healthy levels, the introduced electronegatively charged device filtered particles of a wide range of sizes (10 μm to 1mm) from entering the aortic arch arteries, and with a higher efficacy than for the previous uncharged device. Moreover, the range of filtered particle sizes may be wider than for the only FDA-approved cerebroembolic protection device, the SENTINEL Cerebral Protection System, which has been shown to mainly capture debris of sizes *<* 500 μm [47]. Therefore, the results presented encourage the continuation of the development of this device. The analysis has been conducted on two different patient conditions, healthy and diseased (i.e., suffering from atrial fibrillation) and five different particle sizes have been considered, ranging from 10 μm to 1 mm. A balance was observed between the electrostatic, drag, and inertial forces acting on particles. The particle size was observed to determine which force dominates the particle dynamics. Given the numerical parameters employed in this study, the electrostatic force presents the strongest deflection effect on the smallest particles (10 to 50 μm), which are most effectively diverted by the device. On the other end of the spectrum (500 μm to 1 mm), the electric field cannot overcome the drag and inertial forces, which govern the larger particles’ trajectories, but are mechanically filtered since they cannot fit within the struts. In summary, while the smallest particles are deflected by electrostatic repulsion, the largest ones are mechanically filtered. Therefore, the proposed design effectively blocks all the range of particle sizes analyzed in this study, offering an anticoagulant-free method to prevent stroke and SBIs, especially useful given the growing population of elderly and AF patients. In particular, the results showed that when the device is placed in all three aortic arch arteries, the number of particles entering these arteries was reduced on average by 62.6% and 51.2%, for the healthy and diseased models respectively, matching current oral anticoagulant efficacies. The higher filtering efficiency shown for smallest particles, around 95%, may help prevent microembolisms, associated with accelerated cognitive decline and higher risk of long-term dementia.

## Data Availability

Unfortunately the models and simulation code presented in our paper are not available for public access. Despite our best efforts, we were unable to obtain permission to share the data due to various legal and ethical constraints. Some of the data, however, could be shared upon the request to the corresponding autho

## 7 Future work

The present study presents a methodology to model the problem of interest, and, assuming the considered range of parameters to be physiologic, shows preliminary evidence of the efficacy of the device for thrombus deflection. Nonetheless, further studies will be carried out to improve the solution, verify, and validate the model, validate the ranges of physical parameters employed, and refine the geometrical design for the target application. The scope of the current study was to quantify the efficacy of thrombus deflection by comparing the geometries without deflector vs. with deflector in the three arteries of the aortic arch. This has left the study of other aspects outside the scope of this work. The following are some of the aspects which will be studied in further detail in order to optimize the device design and exhaustively characterize its performance:

- Refine the number of intermediate particle sizes evaluated, between 50 to 500 μm, to determine the critical diameter at which the flow forces dominate over the electrostatic repulsion.
- Guarantee that the parameters employed are within ranges that can assure a sufficiently low patient risk.
- Test the device on heterogeneous populations of virtual patients, that represent real-world diversity, regarding age, sex, and comorbidities.
- Test the electrostatically charged deflector in geometries with different distributions and convexities, respecting the original design of the reference patent ([31]).
- Based on the presented device, a novel one piece design would enable the easy retrieval in TAVR patients 30 − 40 days post-procedure, where in contrast, the design analyzed in the current work, composed of 3 separate deflectors, could be focused on the treatment of elderly patients. The one piece design will be modeled and tested in future iterations.

## 8 Acknowledgements

This research has been supported by “ELEM biotech - the virtual human factory” (www.elem.bio), an INPhINIT fellowship from “la Caixa” Foundation (fellowship ID: LCF/BQ/DI18/11660044), by the European Union’s Horizon 2020 research and innovation program under the Marie Sklodowska-Curie grant agreement No. 713673., by the project CompBioMed2 (H2020-EU.1.4.1.3. Grant No. 823712), by CREXDATA project (HORIZON-CL4-2022-DATA-01. Grant No. 101092749) and by Torres Quevedo Program (PTQ2018-010290), Ministerio de Ciencia e Innovacion, Spain.

### 9 Appendix

**Table 7:**
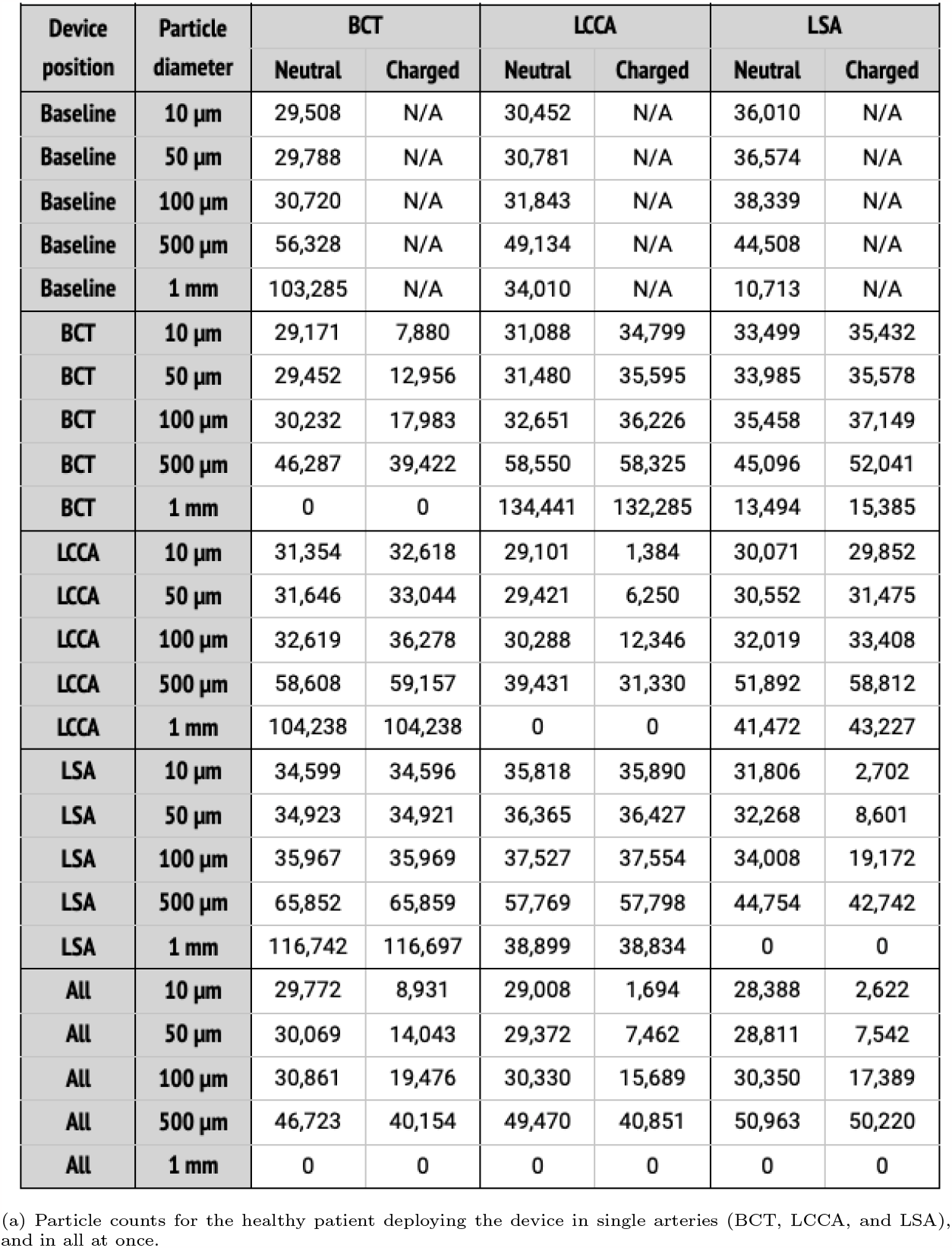
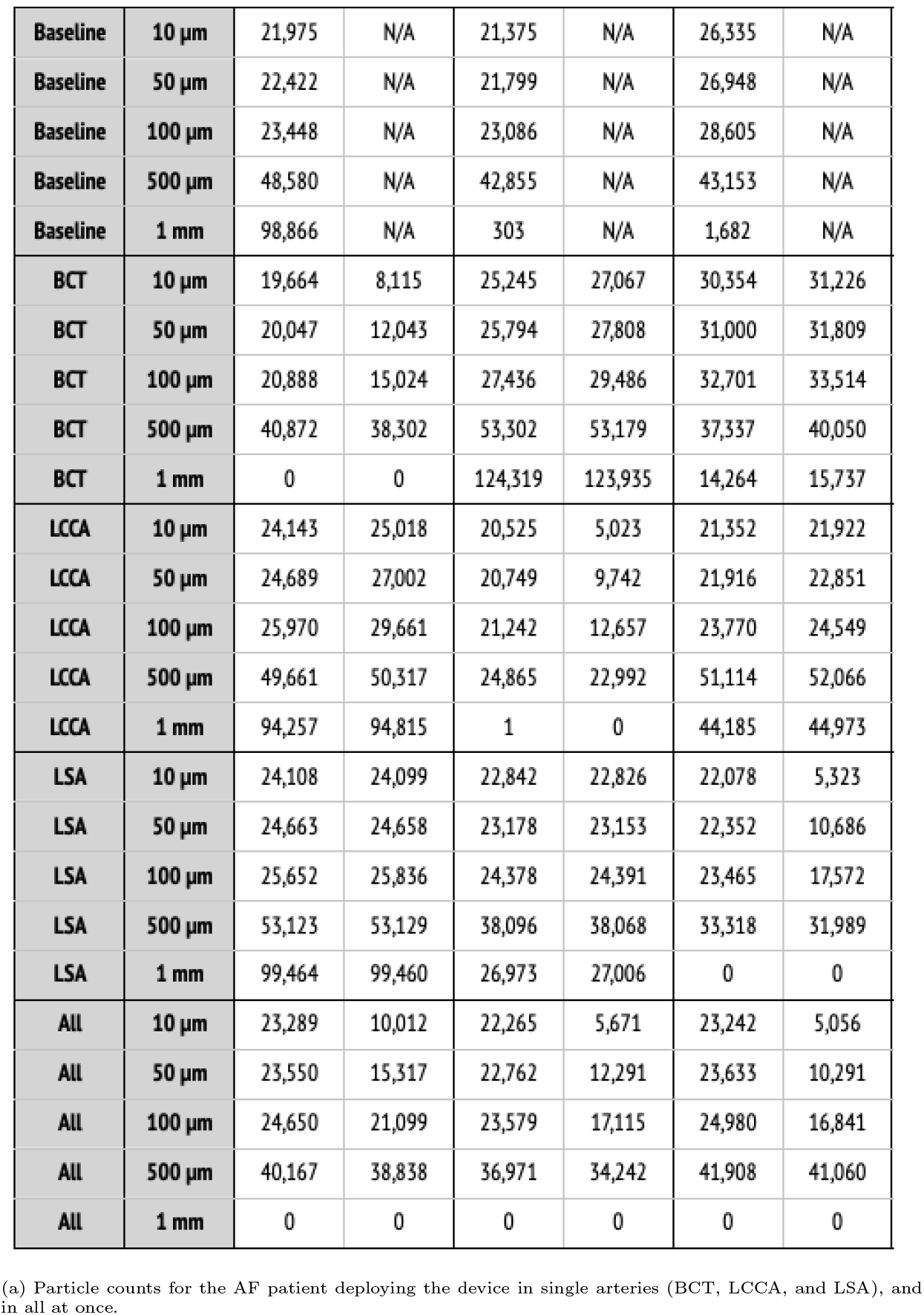
Particle filtering efficacies for the AF patient

| Device position | Particle diameter | BCT |  | LCCA |  | LSA |  |
| --- | --- | --- | --- | --- | --- | --- | --- |
|  |  | Neutral | Charged | Neutral | Charged | Neutral | Charged |
| Baseline | 10 $\mu\text{m}$ | 29,508 | N/A | 30,452 | N/A | 36,010 | N/A |
| Baseline | 50 $\mu\text{m}$ | 29,788 | N/A | 30,781 | N/A | 36,574 | N/A |
| Baseline | 100 $\mu\text{m}$ | 30,720 | N/A | 31,843 | N/A | 38,339 | N/A |
| Baseline | 500 $\mu\text{m}$ | 56,328 | N/A | 49,134 | N/A | 44,508 | N/A |
| Baseline | 1 mm | 103,285 | N/A | 34,010 | N/A | 10,713 | N/A |
| BCT | 10 $\mu\text{m}$ | 29,171 | 7,880 | 31,088 | 34,799 | 33,499 | 35,432 |
| BCT | 50 $\mu\text{m}$ | 29,452 | 12,956 | 31,480 | 35,595 | 33,985 | 35,578 |
| BCT | 100 $\mu\text{m}$ | 30,232 | 17,983 | 32,651 | 36,226 | 35,458 | 37,149 |
| BCT | 500 $\mu\text{m}$ | 46,287 | 39,422 | 58,550 | 58,325 | 45,096 | 52,041 |
| BCT | 1 mm | 0 | 0 | 134,441 | 132,285 | 13,494 | 15,385 |
| LCCA | 10 $\mu\text{m}$ | 31,354 | 32,618 | 29,101 | 1,384 | 30,071 | 29,852 |
| LCCA | 50 $\mu\text{m}$ | 31,646 | 33,044 | 29,421 | 6,250 | 30,552 | 31,475 |
| LCCA | 100 $\mu\text{m}$ | 32,619 | 36,278 | 30,288 | 12,346 | 32,019 | 33,408 |
| LCCA | 500 $\mu\text{m}$ | 58,608 | 59,157 | 39,431 | 31,330 | 51,892 | 58,812 |
| LCCA | 1 mm | 104,238 | 104,238 | 0 | 0 | 41,472 | 43,227 |
| LSA | 10 $\mu\text{m}$ | 34,599 | 34,596 | 35,818 | 35,890 | 31,806 | 2,702 |
| LSA | 50 $\mu\text{m}$ | 34,923 | 34,921 | 36,365 | 36,427 | 32,268 | 8,601 |
| LSA | 100 $\mu\text{m}$ | 35,967 | 35,969 | 37,527 | 37,554 | 34,008 | 19,172 |
| LSA | 500 $\mu\text{m}$ | 65,852 | 65,859 | 57,769 | 57,798 | 44,754 | 42,742 |
| LSA | 1 mm | 116,742 | 116,697 | 38,899 | 38,834 | 0 | 0 |
| All | 10 $\mu\text{m}$ | 29,772 | 8,931 | 29,008 | 1,694 | 28,388 | 2,622 |
| All | 50 $\mu\text{m}$ | 30,069 | 14,043 | 29,372 | 7,462 | 28,811 | 7,542 |
| All | 100 $\mu\text{m}$ | 30,861 | 19,476 | 30,330 | 15,689 | 30,350 | 17,389 |
| All | 500 $\mu\text{m}$ | 46,723 | 40,154 | 49,470 | 40,851 | 50,963 | 50,220 |
| All | 1 mm | 0 | 0 | 0 | 0 | 0 | 0 |
|  |  | Neutral | Charged | Neutral | Charged | Neutral | Charged |
| Baseline | 10 $\mu\text{m}$ | 21,975 | N/A | 21,375 | N/A | 26,335 | N/A |
| Baseline | 50 $\mu\text{m}$ | 22,422 | N/A | 21,799 | N/A | 26,948 | N/A |
| Baseline | 100 $\mu\text{m}$ | 23,448 | N/A | 23,086 | N/A | 28,605 | N/A |
| Baseline | 500 $\mu\text{m}$ | 48,580 | N/A | 42,855 | N/A | 43,153 | N/A |
| Baseline | 1 mm | 98,866 | N/A | 303 | N/A | 1,682 | N/A |
| BCT | 10 $\mu\text{m}$ | 19,664 | 8,115 | 25,245 | 27,067 | 30,354 | 31,226 |
| BCT | 50 $\mu\text{m}$ | 20,047 | 12,043 | 25,794 | 27,808 | 31,000 | 31,809 |
| BCT | 100 $\mu\text{m}$ | 20,888 | 15,024 | 27,436 | 29,486 | 32,701 | 33,514 |
| BCT | 500 $\mu\text{m}$ | 40,872 | 38,302 | 53,302 | 53,179 | 37,337 | 40,050 |
| BCT | 1 mm | 0 | 0 | 124,319 | 123,935 | 14,264 | 15,737 |
| LCCA | 10 $\mu\text{m}$ | 24,143 | 25,018 | 20,525 | 5,023 | 21,352 | 21,922 |
| LCCA | 50 $\mu\text{m}$ | 24,689 | 27,002 | 20,749 | 9,742 | 21,916 | 22,851 |
| LCCA | 100 $\mu\text{m}$ | 25,970 | 29,661 | 21,242 | 12,657 | 23,770 | 24,549 |
| LCCA | 500 $\mu\text{m}$ | 49,661 | 50,317 | 24,865 | 22,992 | 51,114 | 52,066 |
| LCCA | 1 mm | 94,257 | 94,815 | 1 | 0 | 44,185 | 44,973 |
| LSA | 10 $\mu\text{m}$ | 24,108 | 24,099 | 22,842 | 22,826 | 22,078 | 5,323 |
| LSA | 50 $\mu\text{m}$ | 24,663 | 24,658 | 23,178 | 23,153 | 22,352 | 10,686 |
| LSA | 100 $\mu\text{m}$ | 25,652 | 25,836 | 24,378 | 24,391 | 23,465 | 17,572 |
| LSA | 500 $\mu\text{m}$ | 53,123 | 53,129 | 38,096 | 38,068 | 33,318 | 31,989 |
| LSA | 1 mm | 99,464 | 99,460 | 26,973 | 27,006 | 0 | 0 |
| All | 10 $\mu\text{m}$ | 23,289 | 10,012 | 22,265 | 5,671 | 23,242 | 5,056 |
| All | 50 $\mu\text{m}$ | 23,550 | 15,317 | 22,762 | 12,291 | 23,633 | 10,291 |
| All | 100 $\mu\text{m}$ | 24,650 | 21,099 | 23,579 | 17,115 | 24,980 | 16,841 |
| All | 500 $\mu\text{m}$ | 40,167 | 38,838 | 36,971 | 34,242 | 41,908 | 41,060 |
| All | 1 mm | 0 | 0 | 0 | 0 | 0 | 0 |

(a) Particle counts for the AF patient deploying the device in single arteries (BCT, LCCA, and LSA), and in all at once.

